# Epigenetic impairment and blunted transcriptional response to *Mycobacterium tuberculosis* of alveolar macrophages from persons living with HIV

**DOI:** 10.1101/2021.01.26.21250318

**Authors:** Wilian Correa-Macedo, Vinicius M. Fava, Marianna Orlova, Pauline Cassart, Ron Olivenstein, Joaquín Sanz, Anne Dumaine, Renata H.M. Sindeaux, Vania Yotova, Alain Pacis, Josée Girouard, Barbara Kalsdorf, Christoph Lange, Jean-Pierre Routy, Luis B. Barreiro, Erwin Schurr

## Abstract

Persons living with HIV (PLWH) are at increased risk of tuberculosis (TB). HIV-associated TB is often the result of recent infection with *Mycobacterium tuberculosis* (*Mtb*) followed by rapid progression to disease. Alveolar macrophages (AM) are the first cells of the innate immune system that engage *Mtb*, but how HIV and antiretroviral therapy (ART) impact on the anti-mycobacterial response of AM is not known. To investigate the impact of HIV and ART on the transcriptomic and epigenetic response of AM to *Mtb*, we obtained AM by bronchoalveolar lavage from 20 PLWH receiving ART, 16 control subjects who were HIV-free (HC), and 14 subjects who received ART as pre-exposure prophylaxis (PrEP) to prevent HIV infection.

Following *in-vitro* challenge with *Mtb*, AM from each group displayed overlapping but distinct profiles of significantly up- and down-regulated genes in response to *Mtb*. Compared to HC subjects, AM isolated from PLWH and PrEP subjects presented a substantially weaker transcriptional response. Further investigation of chromatin structure revealed that AM from control subjects challenged with *Mtb* responded with pronounced accessibility changes in over ten thousand regions. In stark contrast, AM obtained from PLWH and PrEP subjects displayed no significant changes in their chromatin state in response to *Mtb*. Collectively, these results revealed a previously unknown adverse effect of ART on the epigenetic landscape and transcriptional responsiveness of AM.

## Introduction

Tuberculosis (TB) is globally the leading cause of death due to a single pathogen. In 2018, there were an estimated 1.45 million deaths caused by TB, which included 250,000 people living with the human immunodeficiency virus (HIV) (*1*). Of the estimated 10 million people who fell ill with TB, 850,000 were persons living with HIV (PLWH) indicating a 2.2-fold increased risk of death due to TB in HIV-positive relative to HIV-negative persons. Globally, the proportion of notified HIV-positive TB cases on antiretroviral therapy was 86% with large gaps among countries (*1*). However, even people on long-term antiretroviral therapy (ART) still have significantly increased risk of developing TB (*2*), which is usually the result of new infection rather than reactivation of latent TB (*3-6*).

Transmission of *Mycobacterium tuberculosis* (*Mtb)* occurs by aerosols that are inhaled by an exposed person. Results from the mouse model show that after reaching the lung alveoli, *Mtb* bacilli are being rapidly taken up by alveolar macrophages (AM). Following a delay of approximately 10 days, successful establishment of infection occurs once *Mtb*-infected AM traverse the airway epithelium and establish themselves in the lung interstitium. There, transfer of *Mtb* to permissive inflammatory macrophages occurs and T-cell priming is initiated in the draining lymph nodes (*7*). However, much of the AM-*Mtb* interaction in the alveoli remains unknown.

In humans, not each exposure results in successful infection, and a subset of highly exposed persons entirely resist infection as inferred from the absence of CD4^+^ T cell anti-*Mtb* immunity (*8, 9*). Hence it seems possible that human AM are capable to limit transfer of inhaled *Mtb* bacilli to the lung interstitium. The increased risk of *Mtb* infection and TB disease for PLWH may therefore reflect an impaired ability of AM to restrict *Mtb* to the alveoli, thus placing early events in the *Mtb*-host interaction as a central aspect of TB resistance.

Based on the above considerations, we decided to investigate the transcriptome and epigenome of the *ex-vivo* AM-*Mtb* interaction employing AM obtained from PLWH and HIV-negative participants. Since ART is given to most PLWH, we also included an additional control group consisting of persons who are HIV-negative receiving ART for pre-exposure prophylaxis (PrEP).

## Results

### Study subjects

To investigate the influence of HIV on the ability of AM to mount a response to *Mtb* infection, we studied three groups of subjects: PLWH receiving ART (n=20), persons without HIV but receiving ART as pre-exposure prophylaxis (PrEP; n=14) and persons without HIV not receiving PrEP (healthy controls, HC; n=16, **Table 1**). We obtained AM by BAL from all subjects and challenged the cells *in vitro* with *Mtb*. RNA and DNA were isolated from uninfected and *Mtb* challenged cells and used to assess gene expression levels by RNA-seq, chromatin structure by ATAC-seq, and presence of the histone activation mark H3K27ac by ChIP-seq (**Fig. 1A**). However, due to different quality-control procedures and variable numbers of AM obtained per subject, different counts of subjects were used for the transcriptomics and epigenetics experiments (**Fig. 1A**).

**Table 1.** Demographics and characteristics of enrolled subjects.

| SampleID | Sex | Age at recruitment | Smoker | Recreational drug | Medical history | ART | Prescription drug molecules | Sexually Transmitted Infections | CD4 count |
| --- | --- | --- | --- | --- | --- | --- | --- | --- | --- |
| AMC2 | M | ≥ 40 | YES | Cannabis | None | NO | None | None | 495 |
| AMC7 | M | ≥ 40 | NO | NO | HTN, T1DM, gout | NO | None | None | 500 |
| AMC8 | M | < 40 | NO | NO | None | NO | None | None | 1536 |
| AMC9 | M | ≥ 40 | YES | Cannabis | None | NO | None | None | 1290 |
| AMC10 | M | < 40 | NO | NO | None | NO | None | None | 1091 |
| AMC11 | M | < 40 | NO | NO | None | NO | None | None | 439 |
| AMC12 | M | ≥ 40 | NO | NO | None | NO | None | None | 1263 |
| AMC13 | F | < 40 | NO | NO | None | NO | None | None | 871 |
| AMC14 | F | ≥ 40 | YES | NO | None | NO | None | None | 1023 |
| AMC15 | F | ≥ 40 | NO | NO | None | NO | None | None | 843 |
| AMC16 | F | < 40 | NO | NO | None | NO | None | None | 1249 |
| AMC18 | M | < 40 | NO | NO | None | NO | None | None | 826 |
| AMC19 | M | ≥ 40 | NO | NO | None | NO | None | None | 692 |
| AMC20 | M | < 40 | NO | NO | None | NO | None | None | 545 |
| AMC21 | M | ≥ 40 | NO | NO | HTN, T2DM | NO | None | None | 1077 |
| AMC22 | F | ≥ 40 | YES | Cannabis | None | NO | None | None | 1248 |
| AMP23 | M | ≥ 40 | YES | Cannabis | fibromyalgia | PrEP (2017) | Emtricitabine, Tenofovir_DF | None | 819 |
| AMP25 | M | < 40 | NO | NO | ADHD, depression | PrEP (2016) | Emtricitabine, Tenofovir_DF | None | 575 |
| AMP26 | M | < 40 | NO | Cannabis | chronic.alcoholism | PrEP (2015) | Emtricitabine, Tenofovir_DF | Syphilis | 835 |
| AMP27 | M | < 40 | NO | NO | None | PrEP (2014) | Emtricitabine, Tenofovir_DF | Chlamydia and Gonorrhea | 961 |
| AMP28 | M | ≥ 40 | YES | NO | HTN, T1DM, ADHD | PrEP (2016) | Emtricitabine, Tenofovir_DF | None | 1254 |
| AMP29 | M | < 40 | YES | Cannabis | None | PrEP (2013) | Emtricitabine, Tenofovir_DF | Chlamydia | 981 |
| AMP30 | M | < 40 | NO | NO | depression | PrEP (2016) | Emtricitabine, Tenofovir_DF | None | 847 |
| AMP31 | M | < 40 | NO | NO | None | PrEP (2016) | Emtricitabine, Tenofovir_DF | None | 1163 |
| AMP32 | M | < 40 | NO | NO | None | PrEP (2017) | Emtricitabine, Tenofovir_DF | None | 1069 |
| AMP33 | M | < 40 | NO | Speed | depression | PrEP (2019) | Emtricitabine, Tenofovir_DF | None | 691 |
| AMP34 | M | ≥ 40 | NO | NO | None | PrEP (2016) | Emtricitabine, Tenofovir_DF | Chlamydia | 208 |
| AMP35 | M | < 40 | YES | Cocaine | None | PrEP (2018) | Emtricitabine, Tenofovir_DF | Chlamydia | 832 |
| AMP36 | M | < 40 | YES | Ketamine | None | PrEP (2016) | Emtricitabine, Tenofovir_DF | Gonorrhea | 1009 |
| AMP37 | M | ≥ 40 | NO | Cannabis | None | PrEP (2017) | Emtricitabine, Tenofovir_DF | None | 1454 |
| AMV3 | M | ≥ 40 | NO | NO | HIV (1996) | ART (1998) | Emtricitabine, Tenofovir_DF, Raltegravir | None | 382 |
| AMV4 | M | ≥ 40 | YES | Cannabis | HIV (2002) | ART (2002) | Emtricitabine, Tenofovir_AF, Bictegravir | None | 1172 |
| AMV5 | M | ≥ 40 | NO | NO | HIV (1995) | ART (2009) | Emtricitabine, Tenofovir_DF | None | 606 |
| AMV8 | M | ≥ 40 | NO | NO | HIV (2002) | ART (2013) | Emtricitabine, Tenofovir_DF, Rilpivirine | None | 533 |
| AMV9 | M | ≥ 40 | YES | Cannabis | HIV (2002) | ART (2004) | Emtricitabine, Tenofovir_DF, Raltegravir | None | 536 |
| AMV10 | M | ≥ 40 | NO | NO | HIV (1992) | ART (1996) | Etravirine, Darunavir, Dolutegravir, Ritonavir | None | 269 |
| AMV11 | M | ≥ 40 | YES | NO | HIV (1997) | ART (1999) | Emtricitabine, Tenofovir_DF, Rilpivirine | None | 439 |
| AMV13 | F | ≥ 40 | YES | Cannabis | HIV (1994) | ART (1994) | Emtricitabine, Tenofovir_DF, Rilpivirine, Atazanavir, Raltegravir | None | 716 |
| AMV14 | M | ≥ 40 | YES | Cannabis | HIV (1996) | ART (1997) | Abacavir, Lamivudine, Atazanavir, Ritonavir | None | 983 |
| AMV15 | M | ≥ 40 | NO | NO | HIV (2003) | ART (2004) | Abacavir, Dolutegravir, Lamivudine | None | 468 |
| AMV16 | M | ≥ 40 | YES | NO | HIV (1987) | ART (2002) | Emtricitabine, Tenofovir_AF, Cobicistat, Elvitegravir | None | 1005 |
| AMV17 | M | ≥ 40 | YES | Cannabis | HIV (2014) | ART (2015) | Tenofovir_DF, Dolutegravir, Rilpivirine | None | 1267 |
| AMV18 | M | ≥ 40 | YES | Cannabis | HIV (2016) | ART (2016) | Tenofovir_DF, Dolutegravir, Lamivudine | None | 989 |
| AMV19 | M | ≥ 40 | NO | Cannabis | HIV (2005) | ART (2006) | Abacavir, Dolutegravir, Lamivudine | None | 565 |
| AMV20 | M | ≥ 40 | NO | Cannabis | HIV (1997) | ART (1997) | Emtricitabine, Tenofovir_DF, Cobicistat, Darunavir | None | 727 |
| AMV21 | M | ≥ 40 | YES | Cannabis | HIV (2012) | ART (2012) | Emtricitabine, Tenofovir_DF, Dolutegravir | None | 650 |
| AMV23 | M | < 40 | YES | Cannabis | HIV (2017) | ART (2017) | Emtricitabine, Tenofovir_AF, Cobicistat, Elvitegravir | None | 633 |
| AMV25 | M | ≥ 40 | NO | NO | HIV (2006) | ART (2007) | Emtricitabine, Tenofovir_DF, Efavirenz | None | 906 |
| AMV26 | M | ≥ 40 | NO | NO | HIV (2000) | ART (2010) | Emtricitabine, Tenofovir_DF, Efavirenz | None | 471 |
| AMV27 | M | ≥ 40 | NO | NO | HIV (2017) | ART (2017) | Emtricitabine, Tenofovir_AF, Raltegravir | None | 501 |

Abbreviation list:
**ART**, Antiretroviral Therapy; **HTN**, Hypertension; **T1DM**, Type 1 Diabetes Mellitus; **T2DM**, Type 2 Diabetes Mellitus; **ADHD**, attention deficit/hyperactivity disorder

**Fig. 1.**
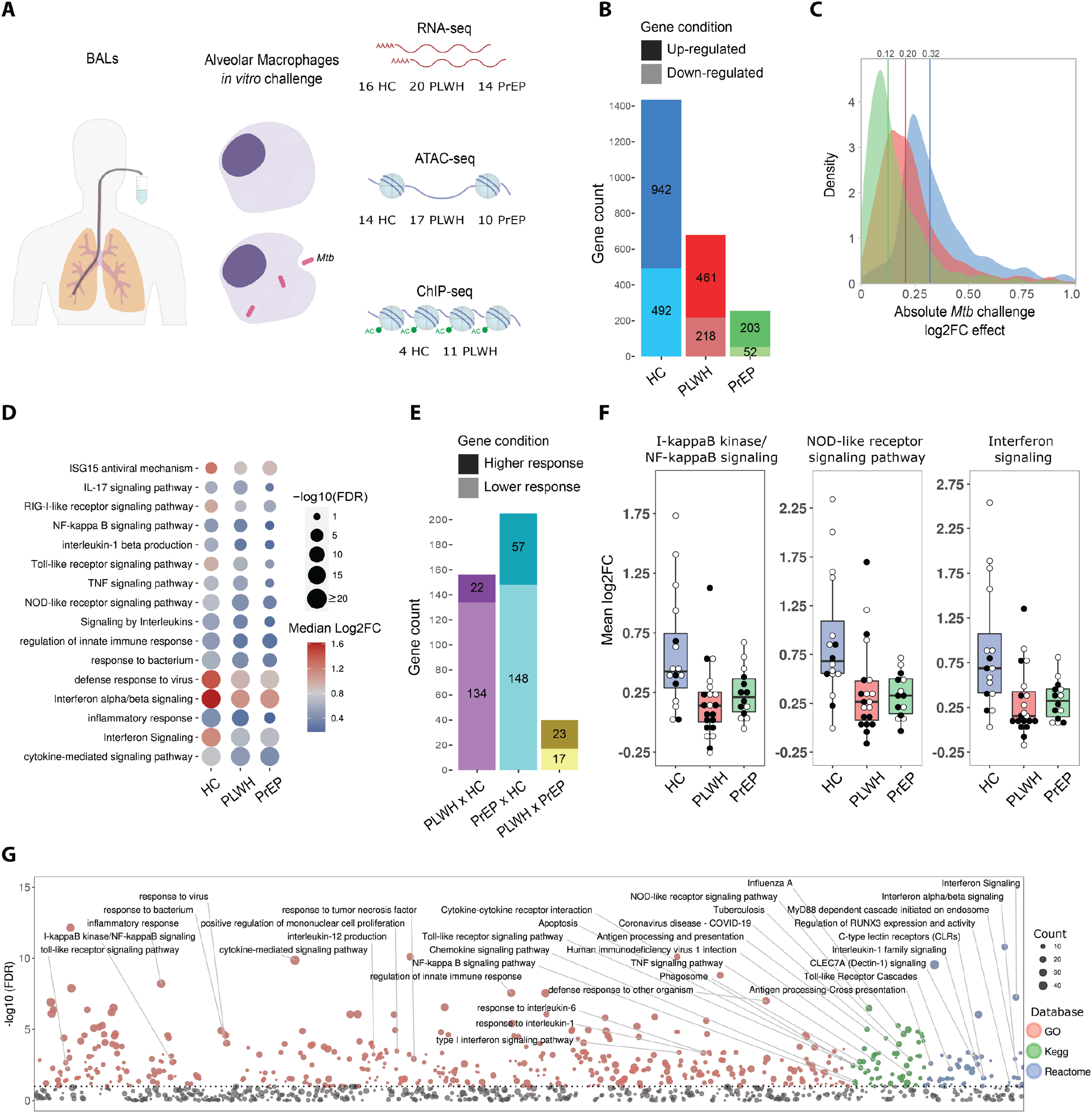
Study design and results for differential gene expression by alveolar macrophages (AM) after challenge with Mycobacterium tuberculosis (Mtb). **(A)** Experimental design and sample number by group. Each subject underwent bronchoalveolar lavage (BAL), AM were obtained from BAL and challenged *in-vitro* with *Mtb*. RNA was obtained for RNA-seq, while DNA was used for ATAC-seq and ChIP-seq experiments. **(B)** Bar graph summarizing differentially expressed genes (DEG) by AM following *Mtb* challenge across the three phenotypic groups (healthy controls – HC; persons living with HIV on antiretroviral therapy – PLWH; subjects on pre-exposure prophylaxis with antiretroviral therapy - PrEP). The y-axis indicates the cumulative DEG count and group ID is indicated below each bar. Dark colour shades represent up-regulated genes (positive log fold-change; logFC) while light shades depict down-regulated genes (negative logFC). **(C)** Density plot presenting logFC as absolute values. All DEGs from panel **B** had their log2FC converted to absolute values and plotted using density function. Vertical coloured lines indicate the median of absolute values per group, with exact values given on top. Blue shade represents HC, red indicates PLWH and PrEP were coloured in green. **(D)** Dot plot presenting significance and overall effect for selected pathways and GO-terms. Pathways/GO-terms are listed on the left, dot sizes reflect the negative log10 FDR values for the enrichment test (bigger dots have smaller *p*-values) and colours from red (higher value) to blue (lower value) indicate the median log2FC across all detected genes in the corresponding pathway or GO term. To derive the average log2FC for a pathway/GO term, for each group we identified the corresponding DEGs, pooled all gene IDs from the three groups, retrieved the log2FC for each of these genes and established the median of log2FC per group. **(E)** Bar graph summarizing genes that displayed significantly different log2FC between groups (differential *Mtb* response). The y-axis indicates the cumulative DEG count while the x-axis indicates the pair-wise group contrast. Dark colour shading represents genes with higher response (e.g. *Mtb* effect in PLWH vs HC subjects resulted in a positive log2FC difference) while light shading depicts genes with lower response (negative log2FC difference). The union of all gene IDs identified across the three contrasts (n=401) results in 333 unique gene IDs. **(F)** Boxplot for three selected pathway/GO terms for genes with significant differential *Mtb* response between PLWH or PrEP against controls (n=302). For each term, the y-axis displays the mean log2FC and dots represent per-subject mean value for pooled DEG. Phenotypic groups are indicated below each box plot across the three terms. White dots represent mean log2FC for subjects who do not smoke cigarettes or used cannabis, while black dots depict smokers and/or cannabis users. To derive the subject mean log2FC, we used pooled DEG from the first two columns in panel E which were significant for interaction contrasts PLWH vs HC and PrEP vs HC, obtained subject-wise log2FC for each DEG and averaged the log2FC from all genes per subject. Results showed that group differences are not driven by outliers and that smoking, or cannabis consumption do not explain group differences. **(G)** Manhattan plot for enrichment test of pathways and GO-terms from merged DEG detected in the differential *Mtb* responses of PLWH vs HC and PrEP vs HC. The y-axis indicates the negative log10 false discovery rate (FDR) values while the tested terms are arranged along the x-axis. The horizontal dashed line represents the 10% FDR cut-off for significant pathways/GO-terms. Dots are sized as a function of the DEG number in a term and colors represent the database from which the terms were obtained. Identified terms represent innate immune processes and intracellular defense mechanisms.

### Alveolar macrophages from PLWH and PrEP subjects display blunted mRNA transcriptomic changes in response to Mtb

Across the three phenotypic groups, we identified 1,626 differentially expressed genes (DEG) in response to *Mtb* (Data File S1). However, there were pronounced DEG count differences in their transcriptional responses (**Fig. 1B**). AM from HC subjects showed 1,434 DEG in response to *Mtb* as compared to only 679 and 255 DEG in PLWH and PrEP subjects, respectively (**Fig. 1B** and Fig. S1A). The magnitude of AM transcriptional response to *Mtb* also differed among the three groups. The smallest mean absolute log2 fold change (log2FC) in response to *Mtb* infection was observed among PrEP subjects while HC subjects displayed the strongest transcriptional response. (**Fig. 1C** and Fig. S1A). To test if *Mtb* effects in PLWH and PrEP subjects were weaker for all DEGs or only for specific gene sets, we assessed the correlations of *Mtb-*triggered logFC across the three groups. We observed that log2FC values were strongly correlated among all three groups, but consistently higher for corresponding genes from HC subjects (Fig. S1B) which suggested a general transcriptional impairment in cells from PLWH and PrEP subjects.

Pathway and GO-term enrichment analysis revealed that *Mtb*-induced transcriptional changes were enriched in biological functions representative of macrophage anti-mycobacterial host responses, such as interferon (IFN) and Toll-like receptor signalling pathways (**Fig. 1D**). However, compared to HC subjects, the number of significant GO-terms/pathways was substantially lower for the PLWH and PrEP groups (Table S1). Moreover, even among shared significant terms, the cumulative transcriptional response of genes in these terms was stronger in the HC group, which further emphasized the blunted anti-mycobacterial response by PLWH and PrEP subjects (**Fig. 1D**). We also tested which genes had a significantly different magnitude of response to *Mtb* among phenotype groups (**Fig. 1E** and Data File S1). We identified 333 genes significantly different between the groups (Data File S2) with the majority displaying lower up-regulation in response to *Mtb* for PLWH and PrEP groups relative to the HC group (Fig. S1, C and D and Table S1). These results provided a formal confirmation that the transcriptional response to *Mtb* challenge by AM of HC was more vigorous compared to the one mounted by PrEP and PLWH subjects.

To explore if the group differences were mainly driven by strong outliers, we derived per subject average log2FC across DEG in significant pathways/GO-terms. We found that AM from HC subjects displayed a more pronounced group transcriptional response to *Mtb*. For example, the mean log2FC for the term “I-kappaB kinase/NF-kappaB signaling” was 3.22-fold (*p* = 2.4×10^−3^) and 2.39-fold (*p* = 3.8×10^−2^) higher in HC as compared to PLWH and PrEP, respectively (**Fig.1F**). GO-terms and pathways tagged by the DEG from PLWH and PrEP vs HC, which were significantly more induced in AM from HC subjects, revealed a predominance of innate-immune and intracellular defense response terms (**Fig. 1G**). This observation alongside the exclusion of possible outlier-driven group differences supported the reduced ability of AM from PLWH and PrEP subjects to mount an effective anti-mycobacterial transcriptional response to challenge with *Mtb*.

### Mtb does not induce chromatin remodelling in AM from PLWH and PrEP subjects

Next, we evaluated *Mtb*-triggered epigenetic changes of AM chromatin accessibility and H3K27 acetylation. Analysis of Differentially Open Chromatin (DOC) in response to *Mtb* indicated that AM from HC had significant increased accessibility (opening) in 8,389 regions and repression (closing) in 3,971 regions (**Fig. 2A**; Data File S3). In contrast, AM isolated from PLWH or PrEP subjects displayed near complete absence of significant chromatin remodeling in response to *Mtb* (**Fig. 2, A and B**). A broad-based epigenetic impairment was confirmed for AM from PLWH subjects by tracking regions with differently H3K27 acetylation (DAc), a mark of active chromatin (**Fig. 3A**; Data File S4). Pathway and GO-term enrichment analyses of genes annotated to DOC and DAc in HC identified typical viral and bacterial response terms (**Fig. 2C**; **Fig. 3B**). As in the transcriptomics analyses, the epigenetic response to *Mtb* by AM from HC strongly implicated host immune response to pathogens and immune-related signalling pathways, which was mainly driven by opening DOCs and regions with increased acetylated H3K27 (**Fig. 2D**; Table S2). Examples of genes with DOCs and DAc at the gene transcription start site (TSS) were *CXCL10, IFI44L*, A*POBEC3A* and *MX1* (Fig. S3 and S4).

**Fig. 2.**
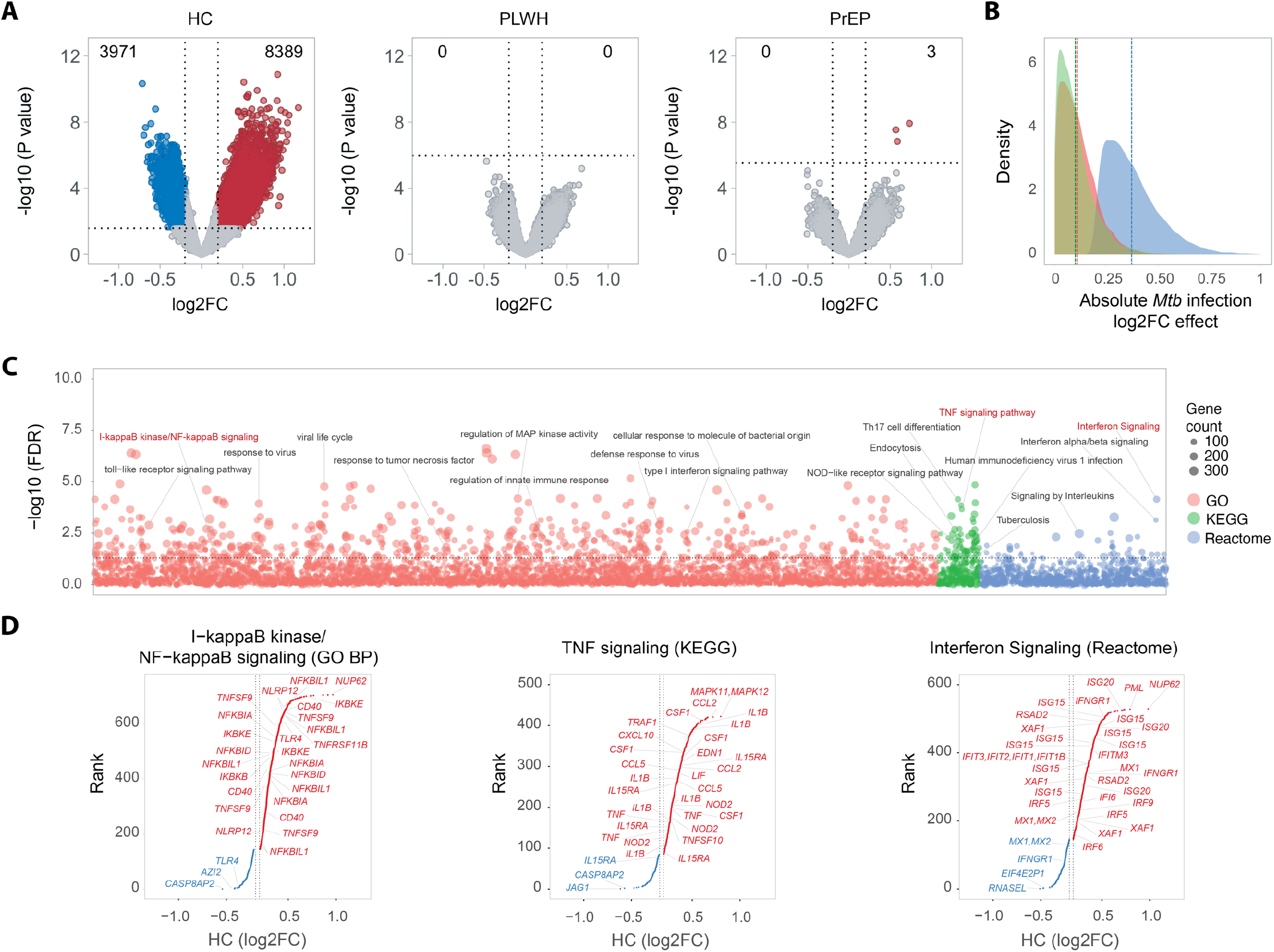
Alveolar macrophage chromatin remodeling in response to Mtb. **(A)** Significance of chromatin changes in response to *Mtb* is plotted against the magnitude of the change. Significantly opened or closed chromatin regions at a false discovery rate (FDR) of 5% after *Mtb* challenge are represented by dots marked in red or blue, respectively. The counts of differential open/closed chromatin (DOC) regions are given at the top left and right corners. Dashed lines indicate the 5% FDR on the y-axis and the minimum log2FC of 0.2 on the x-axis. In healthy controls (HC), 23.3% of the tested regions displayed significant chromatin remodelling in response to *Mtb*, while subjects on pre-exposure prophylaxis (PrEP), and persons living with HIV receiving antiretroviral treatments (PLWH) lacked significant chromatin changes. **(B)** Density of the absolute log2FC effect for 12,360 DOC regions observed in the HC response to *Mtb* is shown for HC, PrEP, and PLWH subjects in blue, green, and red, respectively. The mean log2FC for each group is highlighted by dashed horizontal lines. Both PrEP and PLWH groups show strongly impaired chromatin remodelling in response to *Mtb* compared to HC. **(C)** Pathway/GO-term enrichment analysis of genes assigned to DOC regions for the HC response to *Mtb*. GO biological process (BP), KEGG, and Reactome pathways to which at least five genes had been assigned are plotted against the negative log10 FDR. In total, 724 out of the 8066 tested terms are significant. Enrichment analysis indicated that in AM of HC subjects, *Mtb* challenge promoted chromatin remodelling in regions assigned to genes belonging to IFN and TNF signalling pathways as well as response to viral and bacterial pathogens. **(D)** Rank plot of selected GO BP, KEGG, and Reactome pathways. DOC regions in AM of HC subjects tagged genes assigned to “I−kappaB kinase/NF−kappaB signaling”, “TNF signalling”, and “Interferon signalling” pathways. DOC are plotted according to their log2FC on the x-axis and ranked according to the magnitude of change on the y-axis. Genes corresponding to opening or closing DOC regions are shown in red and blue, respectively. Horizontal doted lines indicate the log2FC cut off of -0.2 and 0.2. In the selected pathways, the majority of the chromatin changes observed indicated increased accessibility in response to *Mtb*.

**Fig. 3.**
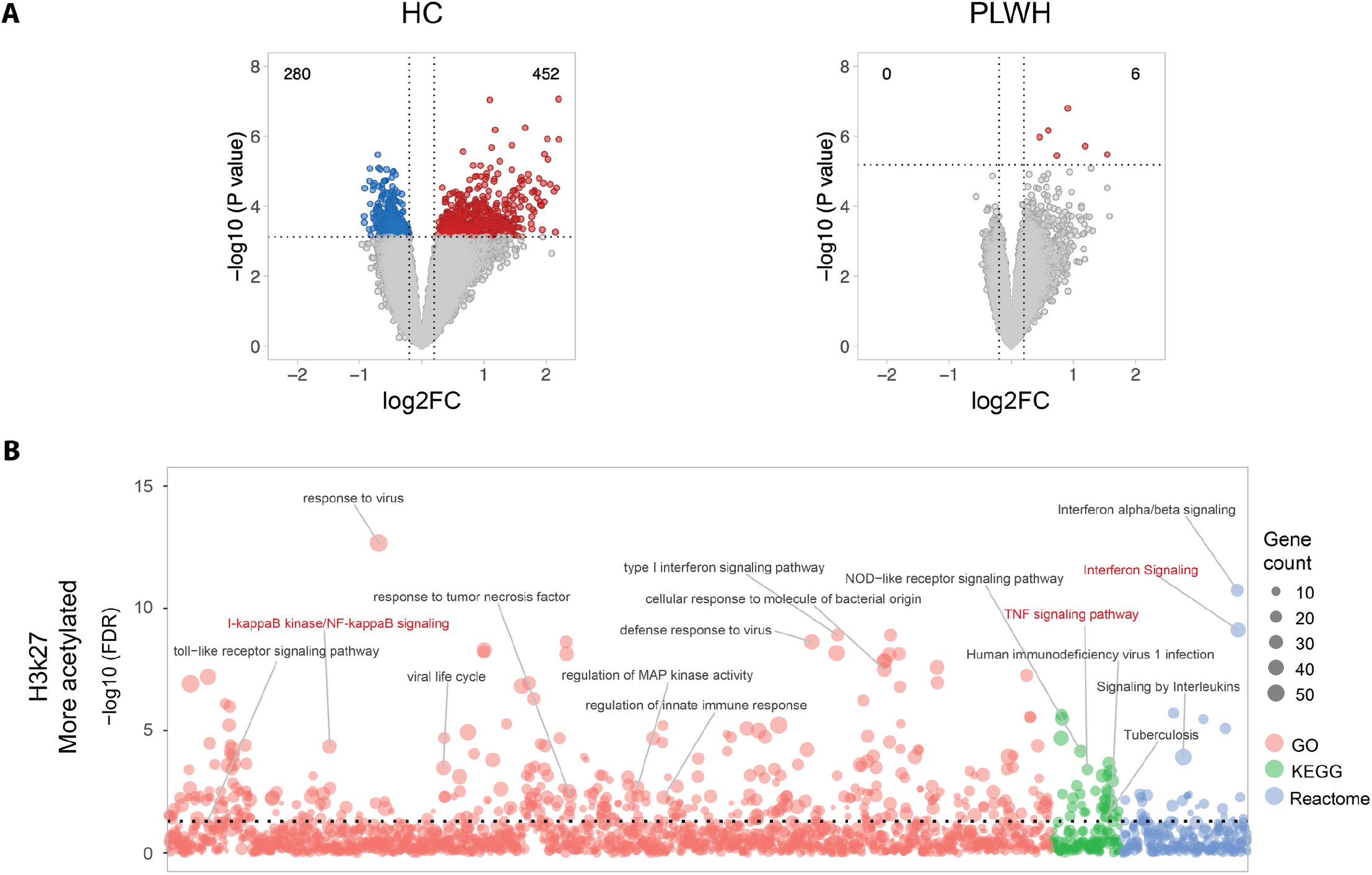
Histone H3K27 acetylation changes in response to Mtb. **a**, Volcano plots for H3K27 acetylation response to *Mtb*. Chromatin regions significantly acetylated at a false discovery rate (FDR) < 5% after *Mtb* challenge are marked in red and blue, respectively. The numbers of acetylated regions are given at the top. Significant changes in H3K27 acetylation, indicating active enhancers, were observed mostly for the healthy control (HC) group, which encompassed five subjects compared to 11 persons living with HIV on antiretroviral therapy (PLWH). While many peaks for H3K27ac did not pass FDR 5%, the magnitude of log2FC was higher in HC compared to PLWH. **b**, Enrichment analysis of genes assigned to regions with increased H3K27 acetylation for the HC response to *Mtb*. GO biological processes (BP), KEGG, and Reactome pathways with at least five assigned genes are plotted against the negative log10 FDR. A total of 167 out of 906 tested terms were significant. As observed for the chromatin changes, enrichment analysis indicated that in AM of HC subjects, *Mtb* challenge promoted higher acetylation in regions assigned to genes belonging to IFN and TNF signalling pathways as well as response to viral and bacterial pathogens.

### Covariates do not explain Mtb-triggered transcriptional and epigenetic differences among groups

The three groups differed with respect to several covariates (**Table 1**). Among the HC group, the proportion of females was higher, and the use of recreational drugs was lower compared to the PLWH and PrEP groups. Subjects in the PrEP group on average were younger compared to the two other groups. The duration of ART was longer for PLWH (mean = 12.12 years, standard deviation (SD) = 7.64) than PrEP subjects (mean = 3 years, SD = 1,47). All PrEP subjects received Emtricitabine and Tenofovir disoproxil fumarate (TDF) while among PLWH there was a wider spread of ART drug regimens. However, only 4 out of 20 subjects had not received TDF at the time of enrolment (**Table 1**). Most of the inter-subject variability was accounted for by the analytical approach employed. Still, given the strong heterogeneity of study subjects we investigated the impact of smoking, recreational drug use and age on *Mtb*-triggered chromatin and transcriptional responses. We found that recreational drug use did not explain the reduced transcriptional activity or absence of chromatin changes in response to *Mtb* among PLWH and PrEP subjects (**Fig. 1F** and **Fig. 4A**). Age and duration of ART/PrEP were also unlikely confounders since PLWH and PrEP subjects differed significantly in age and ART duration, yet both groups showed similar reduced transcriptional activity and lack of chromatin changes relative to HC subjects following challenge with *Mtb* (**Fig. 4, B and C**). Moreover, when stratifying HC and PLWH by age, HC subjects of 40 years and older showed pronounced *Mtb*-triggered chromatin changes that were not observed for PLWH subjects (**Fig. 4D**). In addition to the covariates listed in **Table 1**, we also considered possible group-dependent differences in phagocytic activity. Since the yields of AM were generally not sufficient to conduct independent assessment of phagocytic activity, we used presence of *Mtb* reads in ATAC-seq libraries as proxy and failed to detect any significant group-dependent differences (**Fig. 4E**).

**Fig. 4.**
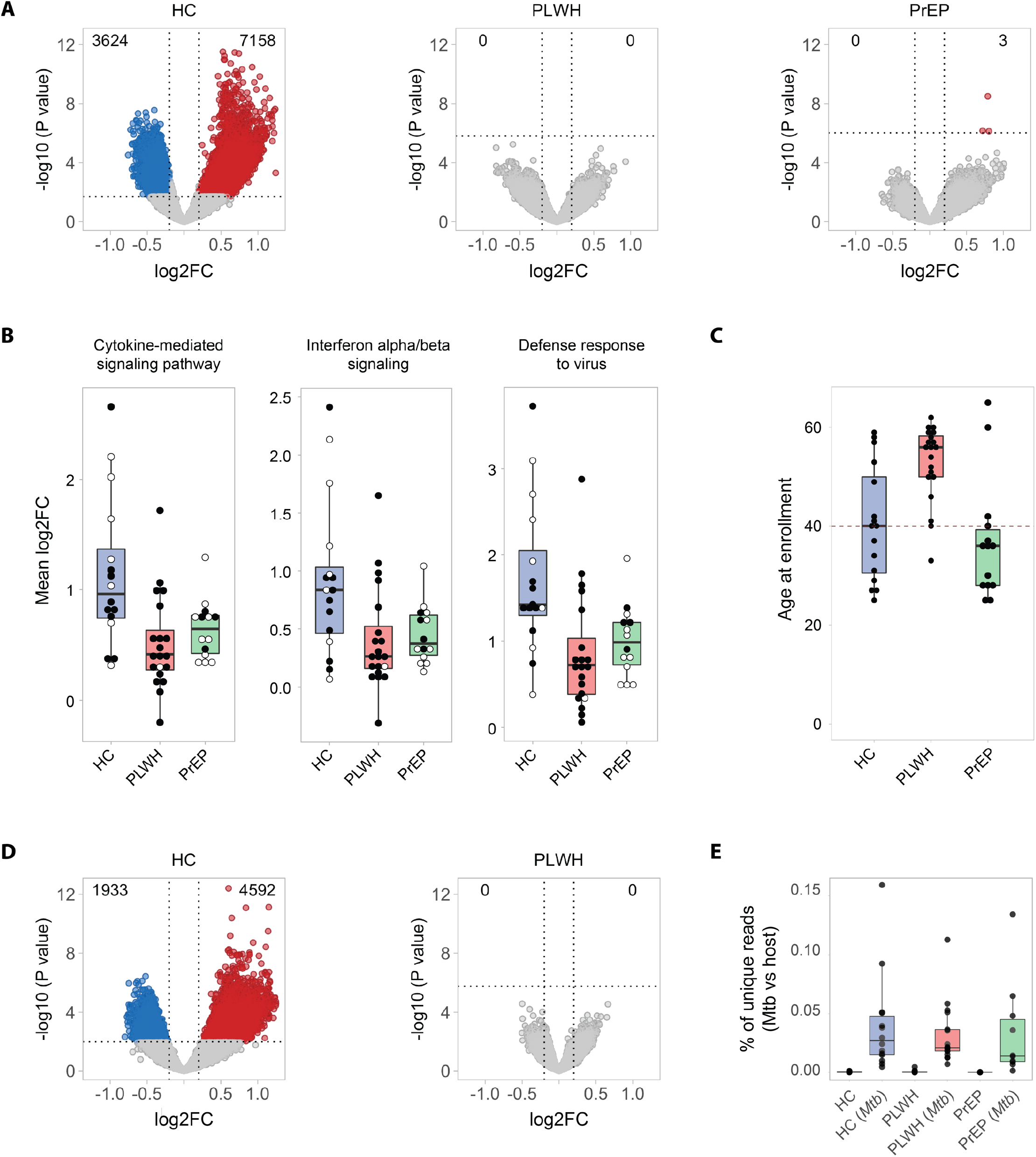
Impact of covariates on Mtb-triggered transcriptional and epigenetic changes in AM. **(A)** Volcano plots for chromatin accessibility changes by AM in response to *Mtb* after removing subjects who smoked cigarettes, cannabis, or both. Included in this analysis were 10 HC, 9 PLWH, and 6 PrEP. Significantly opened or closed chromatin regions at a false discovery rate (FDR) of 5% after *Mtb* challenge are represented by dots marked in red or blue, respectively. The counts of differential open/closed chromatin (DOC) regions are given at the top left and right corners. Dashed lines indicate the 5% FDR on the y-axis and the minimum log2FC of 0.2 on the x-axis. **(B)** Boxplot for three selected pathway/GO terms for genes with significant differential *Mtb* response between PLWH or PrEP against HC (n=302). For each term, the y-axis displays the mean log2FC and dots represent per-subject mean value for pooled DEG from the three groups. White dots represent mean log2FC for subjects younger than 40 years at enrolment, while black dots depict subjects ≥ 40 years of age. To derive the subject mean log2FC, we used DEG from Fig. 1e which were part of significant pathway/GO terms, pooled the genes for each group, obtained the log2FC per subject for each DEG and averaged the log2FC from all genes per subject. **(C)** Boxplot showing the distribution of age at enrolment across the three studied groups. Subjects of the PLWH group were significantly older than the subjects of the healthy controls and PrEP groups. **(D)** Volcano plots for chromatin accessibility changes by AM in response to *Mtb* stratified by age for subjects of the HC and PLWH groups. Subjects tested were ≥ 40 years at enrolment. In this age class the HC, PLWH and PrEP groups encompassed 9, 15 and 3 subjects, respectively. Due to low subject count PrEP subjects were excluded from this stratified analysis. Significantly opened or closed chromatin regions at a false discovery rate (FDR) of 5% after *Mtb* challenge are represented by dots marked in red or blue, respectively. The counts of differential open/closed chromatin (DOC) regions are given at the top left and right corners. Dashed lines indicate the 5% FDR on the y-axis and the minimum log2FC of 0.2 on the x-axis. **(E)** Boxplot of distribution for *Mtb*-aligned reads in ATAC-seq experiments. The y-axis indicates the proportion of total unique paired-end reads aligned to the *Mtb* H37rv genome relative to the Human genome hg38. Unique alignments to *Mtb* genome quantify DNA traces of phagocyted/lysed mycobacteria. In the absence of *Mtb* challenge, little to no alignment was observed to *Mtb* genome. In the challenge condition, the three groups (HC, PLWH, and PrEP) presented a range of proportions of *Mtb* unique reads. No statistical significance was observed between the proportion of *Mtb* reads between the phenotype groups.

### Chromatin accessibility and binding of transcription factors

To link *Mtb*-triggered epigenetic changes in AM from HC subjects with increased gene expression, we evaluated the enrichment of transcription factors (TFs) motifs in *Mtb*-triggered DOC and DAc regions and found an enrichment of IFN-regulatory factor (IRF) motifs in those regions (**Table 2**). Next, we compared the average difference in TF footprint depth between *Mtb* challenged and non-challenge AM to estimate TF activity. Of the 682 TFs obtained from the JASPAR catalog, 21 had significantly higher activity (increased footprint depth) in DOC regions after *Mtb* challenge (**Fig. 5A**). Among these significant TFs were IRF9 (part of the anti-mycobacterial host response) and ZNF684 (a TB biomarker). Of note, *Mtb* challenge had triggered a higher fold change for *IRF9* and *ZNF684* RNA expression in AM from HC subjects compared to AM from PrEP or PLWH subjects (**Fig. 5B**). Moreover, IRF9 and ZNF684 footprints responsive to *Mtb* challenge were enriched in the transcription starting site (TSS) of genes that are part of the IFN signalling pathway (*p* = 8.9 x 10^−6^ and *p* = 0.03, respectively) (**Fig. 5C**). Unexpectedly, we also observed an enrichment of IRF9 active footprints in the TSS of genes of the TNF signalling pathway (*p* = 0.01) (**Fig. 5C**). ZNF684 active footprints were observed in the core promoter of *TNF* after *Mtb* challenge providing a direct link to a major pro-inflammatory cytokine (**Fig. 5D**). Moreover, in support of a coordinated effect of these two TFs, an active footprint for ZNF684 was observed in the promoter region of *IRF9* (**Fig. 5D**).

**Table 2.**
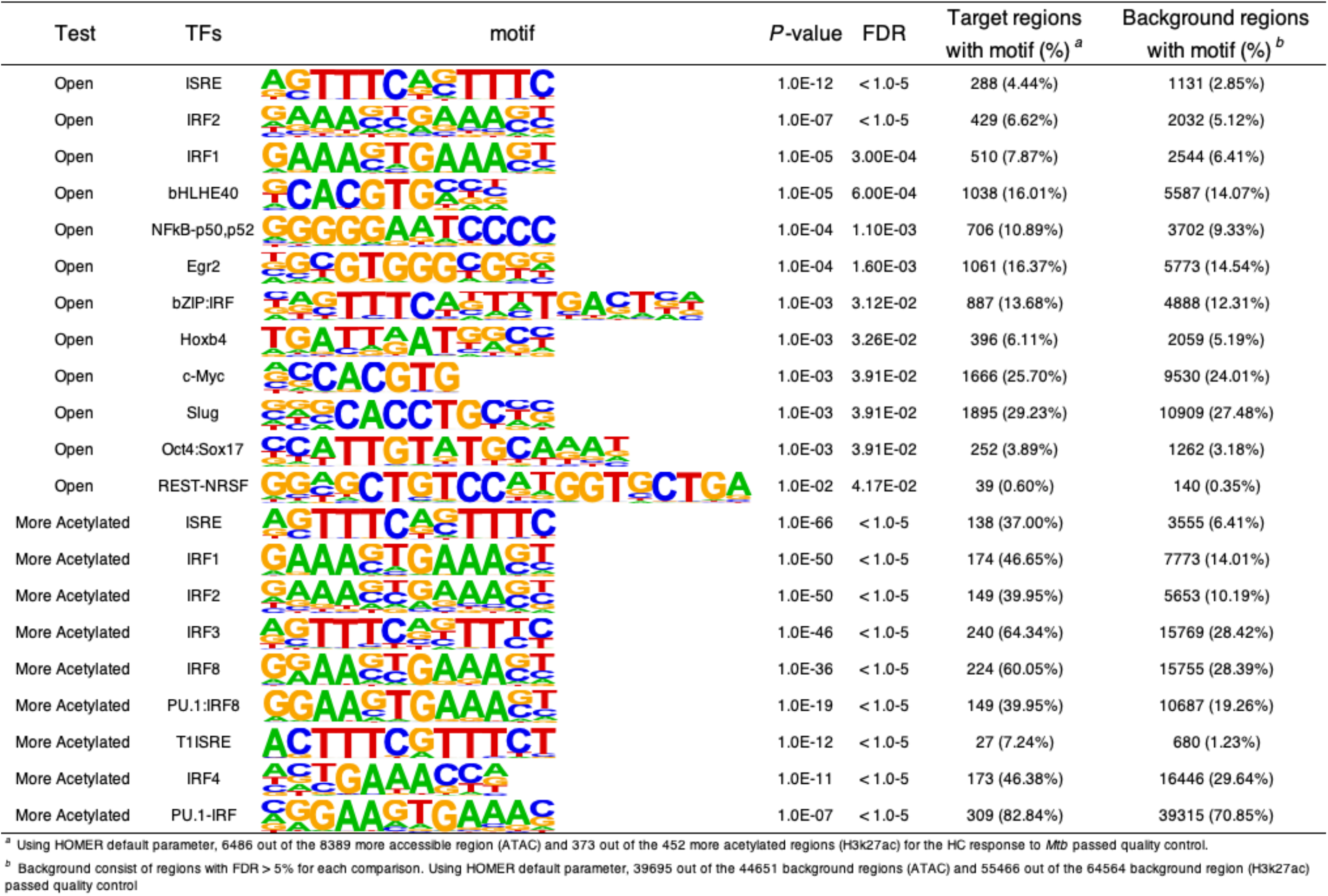
Motif enrichment analysis for ATAC-seq and H3K27ac.

| Test | TFs | motif | *P*-value | FDR | Target regions with motif (%)<sup>a</sup> | Background regions with motif (%)<sup>b</sup> |
| --- | --- | --- | --- | --- | --- | --- |
| Open | ISRE |  | 1.0E-12 | < 1.0-5 | 288 (4.44%) | 1131 (2.85%) |
| Open | IRF2 |  | 1.0E-07 | < 1.0-5 | 429 (6.62%) | 2032 (5.12%) |
| Open | IRF1 |  | 1.0E-05 | 3.00E-04 | 510 (7.87%) | 2544 (6.41%) |
| Open | bHLHE40 |  | 1.0E-05 | 6.00E-04 | 1038 (16.01%) | 5587 (14.07%) |
| Open | NFkB-p50,p52 |  | 1.0E-04 | 1.10E-03 | 706 (10.89%) | 3702 (9.33%) |
| Open | Egr2 |  | 1.0E-04 | 1.60E-03 | 1061 (16.37%) | 5773 (14.54%) |
| Open | bZIP:IRF |  | 1.0E-03 | 3.12E-02 | 887 (13.68%) | 4888 (12.31%) |
| Open | Hoxb4 |  | 1.0E-03 | 3.26E-02 | 396 (6.11%) | 2059 (5.19%) |
| Open | c-Myc |  | 1.0E-03 | 3.91E-02 | 1666 (25.70%) | 9530 (24.01%) |
| Open | Slug |  | 1.0E-03 | 3.91E-02 | 1895 (29.23%) | 10909 (27.48%) |
| Open | Oct4:Sox17 |  | 1.0E-03 | 3.91E-02 | 252 (3.89%) | 1262 (3.18%) |
| Open | REST-NRSF |  | 1.0E-02 | 4.17E-02 | 39 (0.60%) | 140 (0.35%) |
| More Acetylated | ISRE |  | 1.0E-66 | < 1.0-5 | 138 (37.00%) | 3555 (6.41%) |
| More Acetylated | IRF1 |  | 1.0E-50 | < 1.0-5 | 174 (46.65%) | 7773 (14.01%) |
| More Acetylated | IRF2 |  | 1.0E-50 | < 1.0-5 | 149 (39.95%) | 5653 (10.19%) |
| More Acetylated | IRF3 |  | 1.0E-46 | < 1.0-5 | 240 (64.34%) | 15769 (28.42%) |
| More Acetylated | IRF8 |  | 1.0E-36 | < 1.0-5 | 224 (60.05%) | 15755 (28.39%) |
| More Acetylated | PU.1:IRF8 |  | 1.0E-19 | < 1.0-5 | 149 (39.95%) | 10687 (19.26%) |
| More Acetylated | T1ISRE |  | 1.0E-12 | < 1.0-5 | 27 (7.24%) | 680 (1.23%) |
| More Acetylated | IRF4 |  | 1.0E-11 | < 1.0-5 | 173 (46.38%) | 16446 (29.64%) |
| More Acetylated | PU.1-IRF |  | 1.0E-07 | < 1.0-5 | 309 (82.84%) | 39315 (70.85%) |

<sup>a</sup> Using HOMER default parameter, 6486 out of the 8389 more accessible region (ATAC) and 373 out of the 452 more acetylated regions (H3K27ac) for the HC response to *Mtb* passed quality control.
<sup>b</sup> Background consist of regions with FDR > 5% for each comparison. Using HOMER default parameter, 39695 out of the 44651 background regions (ATAC) and 55466 out of the 64564 background region (H3K27ac) passed quality control

**Fig. 5.**
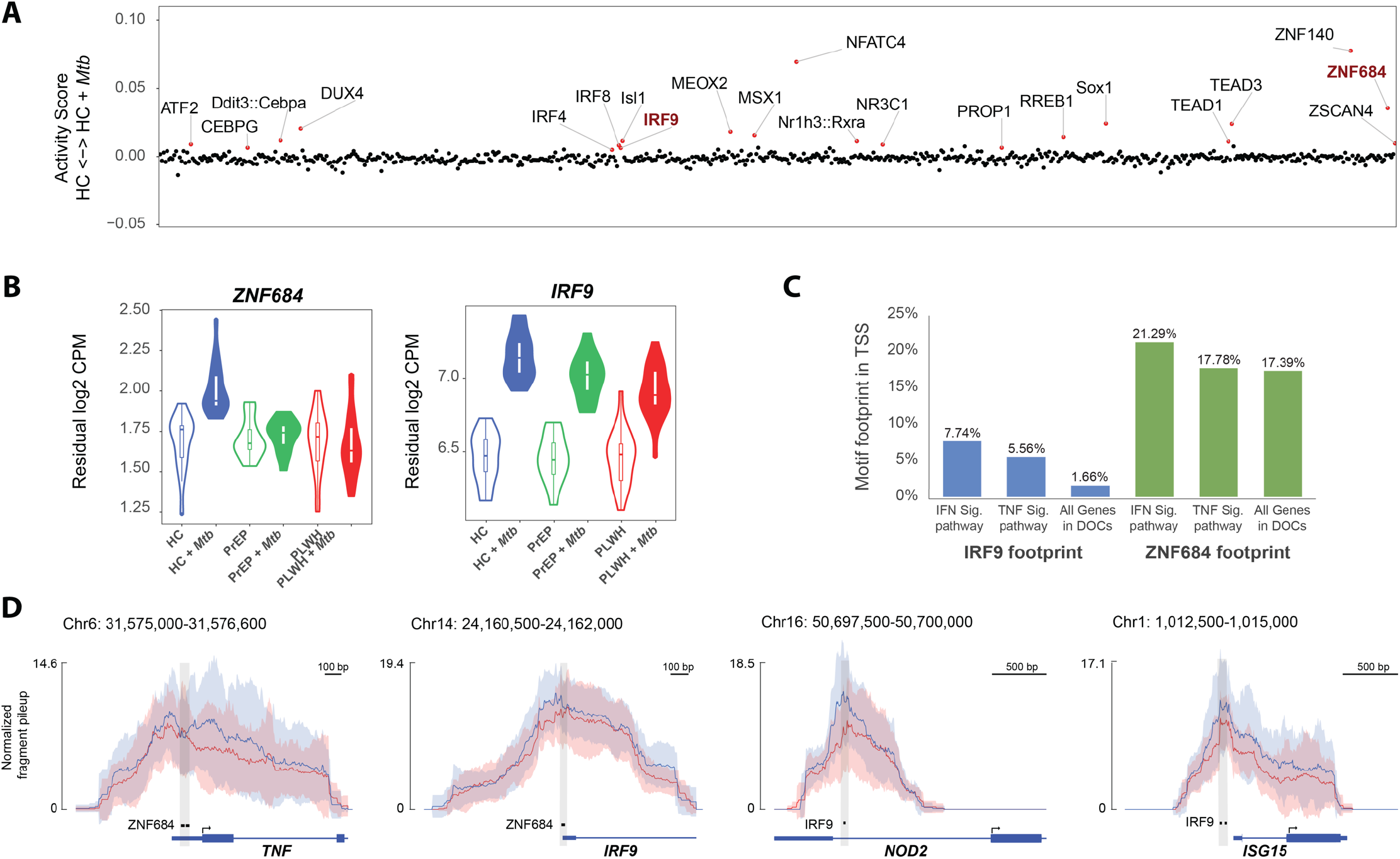
Transcription factor (TF) footprint activity in differential open/closed chromatin (DOC) regions. **(A)** Activity scores denoting differences in depth of TF footprints between healthy controls (HC) and HC + *Mtb* AM are plotted on the y-axis for 682 TFs from the JASPAR catalog showing a minimum of 50 footprints in DOC regions. A positive activity score indicates that footprints for a TF are more pronounced in HC after *Mtb* infection. Twenty-one TF with significant activity score at FDR < 5% are displayed as red dots. **(B)** mRNA expression levels for the transcription factors IRF9 and ZNF684 are shown as violin plots. The residual log2 copy per million (CPM) reads, after removing inter-individual variability, is plotted for each group i.e., healthy controls (HC), subjects on pre-exposure prophylaxis (PrEP), and persons living with HIV receiving antiretroviral treatments (PLWH) and condition (*Mtb*-challenged or not). **(C)** Percentage of genes in the TNF or IFN pathways that have IRF9 or ZNF684 footprints in their transcription starting site (TSS; ± 5kb from the promoter) was compared to all genes with TSS in DOC regions. As expected, we observed that IRF9 footprints are more frequent in genes assigned to Reactome’s IFN signalling pathway compared to random TSS in DOCs. Surprisingly, IRF9 footprints were also more frequent than by chance in KEGG’s TNF pathway. **(D)** ZNF684 and IRF9 footprints in selected IFN and TNF genes. The mean normalized fragment pileup is plotted on the y-axis as red and blue lines for non-infected and *Mtb*-challenged AM from HC, respectively. The standard deviation of the means is shown by shades for each condition. The location of the motif assigned to the footprint is shown by a black bar at the bottom. We detected a pronounced footprint for ZNF684 in the core promoter of the *TNF* gene and also in the promoter of *IRF9*. Motifs for IRF9 were detected in footprints located close to the promoter of the *NOD2* and *ISG15* genes.

To further assess the link between *Mtb*-dependent TF binding and RNA expression, we evaluated if the 135 expressed genes with active IRF9 footprint at the TSS were differentially expressed in HC following *Mtb* challenge. We observed that 65 out of the 135 genes were DEGs (57 up-regulated and 8 down-regulated) following *Mtb* challenge (*p* = 7.9 x 10^−23^, Table S3 and Data File 5). AM from PLWH and PrEP had lower logFC in response to *Mtb* for all 65 DEGs (Data File 5). Of the 57 up-regulated DEGs with IRF9 TSS footprint, 17 were part of the IFN or TNF pathways, including *NOD2* and *ISG15* (**Fig. 5D**). Collectively, these data showed that the epigenetic response observed in AM from HC subjects promoted the binding of key transcriptions factors required for IFN and TNF signalling. The locked state of chromatin in AM from PLWH and PrEP subjects was therefore likely to account, at least in part, for the reduced transcriptional activity of those cells in response to *Mtb*.

## Discussion

There is a scarcity of knowledge about the early fate of inhaled *Mtb* bacilli in the human lung. Infection of human hosts with *Mtb* is inferred from detectable anti-*Mtb* immunity through either the *in-vivo* tuberculin skin test (TST) or *ex-vivo* IFN-γ release assays (IGRAs) (*10*). However, these tests reflect T-cell immunity that is being generated in the lymph nodes, and not in the lung, several weeks after the initial uptake of *Mtb* (*11*). Consequently, much of our knowledge about the *Mtb*-host interplay from inhalation to the development of acquired T cell immunity has been derived from observations in animal models. In the mouse, *Mtb* bacilli in the lung alveoli are taken up by AM (*7*). Infected AM function as niche for *Mtb* replication prior to the migration of *Mtb-*infected AM to the lung interstitium where *Mtb* are transferred to inflammatory macrophages that facilitate the induction of *Mtb*-specific T cell immunity (*7*). The increased permissiveness of AM for *Mtb* in the mouse has been confirmed in a number of independent studies and appears to reflect the metabolic state of the cells (*12, 13*).

It is not known how well this scenario is replicated in the human lung. One critical difference between the mouse model and humans is the ability of a subgroup of *Mtb*-exposed persons to avoid the development of classical T-cell immunity and TB disease even in the presence of long lasting *Mtb* exposure (*9*). Recently, the phenotype of strong resistance to immune conversion of persons living in high *Mtb* exposure settings has been confirmed for HIV-infected but immune-reconstituted people (*14*). While the underlying mechanisms that preclude the development of classical T cell immunity are unknown, initial results implicate both non-classical T cells and B cells (*14, 15*). How and where AM interact with these cells of the acquired immunity arm is not known. We also do not know how the strong persistent exposure of human AM to environmental stimuli and pollutants impacts on AM physiology and anti-mycobacterial activity. An important influence of the environment on AM physiology is consistent with the pronounced inter-individual differences observed in our study. Nevertheless, in our experiments all AM displayed an innate anti-mycobacterial immune response to *Mtb* that was dominated by IFN and TNF signalling suggesting that AM are directly involved in the host defense against *Mtb*. However, this anti-*Mtb* response was significantly weaker in AM from PLWH and PrEP subjects.

The impetus for our experiments was the well-known observation that PLWH are at increased risk to become infected with *Mtb* and to develop clinical TB disease (*4-6, 16*). This increased susceptibility is still present in PLWH who have been immune–reconstituted with ART (*2, 17*). A detrimental effect of HIV on AM function has been shown in pre-ART patients including on their response to *Mtb* (*18*). In our study we found no evidence for reduced phagocytic uptake of *Mtb* by AM across phenotypic groups. This is consistent with previous reports that AM obtained from PLWH showed no defect in phagocytosis of *Salmonella Typhimurium* compared to HIV-negative controls (*19*). AM from PLWH on long term ART displayed a decreased ability to kill *Streptococcus pneumoniae* due to their HIV gp120-mediated reduced AM apoptotic activity (*20, 21*). Consistent with this finding, reduced AM phagocytic activity of *Staphylococcus aureus* in a sample of PLWH/ART subjects was associated with detectable proviral DNA (*22*). In our sample we found that 6 out of the 7 tested PLWH subjects had undetectable proviral counts arguing against a role of provirus in our sample (*23*). Moreover, the differences in the transcriptional response to *Mtb* between the PrEP and PLWH groups were less pronounced as compared to the differences of both groups to HC participants which argues against HIV being the main cause of the blunted anti-*Mtb* response by AM.

The differences in the transcriptional response between the phenotype groups were graded and quantitative. However, the changes in *Mtb*-triggered chromatin structure of AM were sharply divided between PLWH and PrEP subjects compared to controls. Indeed, AM from both PLWH receiving ART and PrEP subjects displayed a near total lack of chromatin changes in response to *Mtb*. This suggested an unknown effect on chromatin of the drugs used in the PrEP and ART regimen. Yet, how drugs might impact the AM epigenetic responsiveness and the transcriptional response to *Mtb* is not clear. While the observed increased TF binding in DOC/DAc regions is consistent with the idea of the chromatin lock-down being the cause of the reduced transcriptional response, this causality remains to be more stringently established. Irrespective of the precise mechanisms, the blunted ability of AM to mount a response to an infectious insult may contribute to the known increased vulnerability to pulmonary disease of PLWH. For example, the prevalence of bacterial pneumonias and chronic obstructive pulmonary disease (COPD) are significantly increased in PLWH/ART patients (*24, 25*). PLWH/ART patients also experience an increased risk of sepsis with poor outcome (*26*). However, a more direct link of our findings can be made with TB for which PLWH on long term ART who have been immune– reconstituted remain at increased risk (*2, 17*). Since our data suggest that the blunted AM response to *Mtb* is predominantly mediated by the anti-viral drugs and not the virus, use of PrEP in settings of high TB transmission might require to consider the risk of TB versus protection from HIV. To our knowledge, no data are presently available that link PrEP with risk of TB. We note, however, that PrEP has been associated with reduced risk of SARS-CoV-2 virus infection in Spain (*27, 28*). Efficient infection of epithelial cells by SARS-CoV-2 is dependent on the virus induced up-regulation of its ACE2 receptor (*29*). If the impact of PrEP on chromatin structure described here for AM also hold true for epithelial cells, this may underlie the protective effect observed.

## Materials and Methods

### Subjects

The study was approved by the Research Ethics Board of the McGill University Health Center (MUHC) (MP-CUSM-15-406). Participants were recruited at the McGill University Health Centre (MUHC, Montreal, Canada). HIV negative participants represented a control group (n=23) and a high-risk group on preventive therapy (PrEP, n=14). HIV positive participants (PLWH, n=27) were receiving antiretroviral therapy (ART) to suppress peripheral viral load for at least 3 years and without any active respiratory symptoms or infections. Exclusion criteria included past history of TB, pregnancy, chronic cardiovascular or active pulmonary infection and immunosuppressive medication. All participants were tested with QuantiFERON Gold, Cat#0594-2010/Cat#T0590-0301 (Qiagen, Germany) and one subject each of the HC and PrEP groups tested positive. Since exclusion of these two subjects had no significant impact on reported results, we left them included in the present presentation of the data. Information on participant characteristics and antiretroviral drugs was extracted from the clinical database and participant electronic records. Of these enrolled subjects, we excluded 7 HC and 7 PLWH from the analyses due to low yield of BAL cells, or inability to pass quality control for sample or library preparation.

### Bronchoalveolar lavage and sample processing

Bronchoscopies were performed at the Centre for Innovative Medicine of the Research Institute of the MUHC following the guidelines of the American Thoracic Society (*30*). Briefly, standard flexible bronchoscopy of the middle lobe was conducted under local anesthesia with lidocaine with additional conscious sedation with intravenous midazolam. A total of 200 ml sterile saline instilled in 50-ml volumes resulted in an average return of 115 ml (range of 50-150 ml) of bronchoalveolar lavage (BAL) fluid. BAL fluid in non-adherent tubes was stored on ice for a maximum of 30 min prior to the isolation of AM. All BAL fluids were strained through a 100μm filter to remove any mucus and cell clumps. Cytology assessment of BAL samples indicated that AM represented >90% of all recovered mononuclear cells.

### Alveolar macrophage preparation

AM were isolated by adhesion. Collected BAL cells were spun for 10 minutes at 300g, 10°C, and washed twice in RPMI-1640 with L-glutamine (Wisent, Canada), containing 2% human serum (heat-inactivated AB+ off the clot, Wisent, Canada), 1% penicillin/streptomycin (Gibco, USA), 10mM HEPES (Gibco, USA), 1% Non-essential amino acids (Gibco, USA) and 1.75ng/ml Amphotericin B (Wisent, Canada). After the cell count, 1.5×10^6^ AM were seeded per well of a 6-well plate (Falcon, USA) in RPMI-1640 supplemented with 10% human serum and incubated for 1.5h-2h at 37°C, 5% CO2, and 95% relative humidity. Non-adherent cells were removed by thorough washings with pre-warmed (37°C) RPMI-1640 supplemented with 2% human serum. Washed adherent AM were then infected with freshly prepared *Mtb*. An estimate for the number of adherent AM was derived by subtracting the number of detached from the number of seeded cells.

### Mycobacterial cultures and macrophage challenge in vitro

Virulent *Mtb* strain H37Rv was grown in a liquid culture of Middlebrook 7H9 medium (Difco, Detroit, USA, #295939) containing 0.2% glycerol (Fisher), 0.05% Tween-80 (Sigma-Aldrich) and 10% albumin-dextrose-catalase (ACD, Fisher) at 37°C in rolling incubators. Bacteria were grown to log phase, as determined by an optical density of 0.3 to 0.8 at 600nm, prior to inoculum preparation. Further, bacterial cultures were spun for 15 minutes at 3700 rpm, resuspended in RPMI-1640 and dislodged with a 22G needle. Cell suspensions were filtered through 5μm filters (Millipore, USA) to ensure single mycobacteria suspensions for the challenge experiments. Bacterial counts of inocula were done using disposable Neubauer hemocytometer (C-Chip, INCYTO, South Korea). Bacterial loads were confirmed by colony-forming unit (CFU) counts by plating serial dilution of inoculum in 7H9 growth medium on Middlebrook 7H10 agar (Becton Dickinson, Franklin Lakes, NJ, USA, #254520) plates containing 0.5% glycerol (Fisher) and 10% oleic acid-albumin-dextrose-catalase (OADC, Becton Dickinson, Franklin Lakes, USA, #211886). Colonies were counted 3-4 weeks post-plating. AM were infected at a multiplicity of infection (MOI) of 5:1 for 18-20hrs 37°C, 5% CO2, and 95% relative humidity.

### Preparation of RNASeq and ATACSeq libraries

For downstream RNA and ATAC experiments AM were rinsed with warm PBS to remove damaged cells and debris before further processing. For RNA, cells were lysed with Trizol and kept at -80°C until extraction. For RNA isolation the miRNeasy kit (Qiagen, Germany) was used and RNA integrity (RIN) was assessed with the Agilent 2100 Bioanalyzer (Agilent Technologies, Waldbronn, Germany). Only samples with RIN >8 were selected for library preparation using TruSeq RNA Library Preparation Kit v2, Set A (Illumina, USA).

For chromatin accessibility using the ATACseq method (*31, 32*); AM were lifted with pre-warmed CellStripper solution (Corning, USA) and gentle pipetting. Viability was assessed with trypan blue staining, and only samples with viability > 85% were used. We followed the protocol by Buenrostro *et al*. (*32*). Briefly, 50,000 cells were used for generating challenged and non-challenged libraries. AM were permeabilized with lysis buffer containing 0.05% IGEPAL. Nuclei were incubated with Tn5 transposase (Illumina, USA) and further ATAC-seq libraries were amplified for a total of 5-10 PCR cycles. Barcoded and multiplexed RNA or ATAC-seq libraries were sequenced on an Illumina HiSeq4000 sequencer at Genome Quebec, Montreal, Canada.

### ChIP-seq library preparation and sequencing

Samples from challenged and non-challenged AM were crosslinked with 1% w/v formaldehyde for 10 min at RT and immediately quenched for 5 min with 125 mM Glycine at RT. The formaldehyde fixed samples were then sonicated to 150-500 bp using a S220 (Covaris) and then ChIP-DNA prepared using a manual chromatin immunoprecipitation method with an Antibody-Antigen incubation of 18 hrs., followed by bead incubation for 135 minutes, and 6 x 5-minutes washing steps.

Approximately 0.5 million cells were used for each ChIP and ∼50,000 cells for the input. We used H3K27ac antibody (Abcam, Ab4729, GR3211959-1). ChIP and Input libraries were prepared using the MicroPlex Library Preparation Kit (C05010012, Diagenode), with alterations including PCR enrichment (7 to 20 cycles) prior to size selection and use of Ampure beads for size selection (250-350 bp). Libraries for ChIPSeq were sequenced on an Illumina NovaSeq 6000 S2 SR100.

### Data processing and statistical analysis

Detailed description for raw sequences processing to statistical analyses are provided in Supplementary Information. Briefly, Baseline and challenged transcriptome and epigenomic landscapes of AMs were determined following published protocols (*31, 33*). Gene expression readouts were generated using alignments and annotated set of transcripts as input to Salmon (*34*). Differences in chromatin accessibility and H3K27ac levels were assessed with a count-based peak quantification as shown in (*35*). Quantification files from each approach were merged as a single matrix, genes and peaks with low counts were filtered out and libraries were normalized, scaled and log2 transformed using edgeR and Limma-voom (*36, 37*).

For the statistical analysis, we defined linear models with blocking design on subject ID to account for inter-individual differences and modelled the challenge effect nested within phenotypic groups. For all experiments we tested the challenge effect for HC, PrEP and PLWH subjects, while for RNAseq we also contrasted the phenotype groups against each other (interaction test). ATACseq and ChIPseq p-values were adjusted with Storey’s false discovery rate (FDR) and peaks considered significant if FDR < 0.05 and absolute log fold-change (logFC) > 0.2. These were termed differentially open chromatin (DOC) for ATACseq and differentially acetylated (DAc) when referring to ChIPseq. For RNA-seq we employed a two-stage FDR procedure and we considered genes to be differentially expressed genes (DEG), if absolute logFC ≥ 0.2 and stageR *p*-val ≤ 0.05 (*38*).

We performed pathway and gene ontology enrichment analysis using three independent databases (KEGG, Reactome and AmiGO). Enrichment results were merged and the Benjamini-Hochberg FDR method was applied to raw p-values. We tested TF motif enrichment in DOCs and DAc regions using HOMER (*39*). *Mtb*-triggered differences in TF footprint activity over DOC regions were estimated with HINT-ATAC (*40*). TF motifs with FDR < 0.05 and activity score > 0.005 were considered significant.

## Data Availability

All data associated with this study are present in the main text or the Supplementary Materials. RNA, ATAC and ChIP-seq raw sequences and corresponding quantification matrices analysed is this work were submitted to NCBI's Gene Expression Omnibus repository (data is being processed, accession number will be available for publication).

## Acknowledgments

We thank all volunteers who participated in this study. We thank Pilar Domenech for help with *Mtb* cultures. We thank personnel of the CL3 platform of the RI-MUHC and the Centre d’expertise et de services Génome Québec. We thank all members of the Schurr and Barreiro labs for discussions of the data and helpful comments on the manuscript.

## Author Contributions

W.C.-M. analysed the transcriptomic data and V.M.F. analysed the epigenomic data. E.S., L.B.B., W.C.-M. and V.M.F designed the study. M.O. and P.C. prepared samples, and performed cell-based experiments for RNA-seq, ATAC-seq and ChIP-seq. A.D., R.H.M.S. and V.Y. performed ChIP experiments. J.-P.R. and J.G enrolled subjects. R.O. performed bronchoalveolar lavages. J.S. and A.P. provided analytical guidance and participated in initial data analyses. B.K. and C.L. provided experimental guidance for AM isolation from BAL samples. E.S. and L.B.B. procured funding sources W.C.-M., V.M.F., L.B.B. and E.S. wrote the draft of the manuscript. All authors participated in the final assemble of the manuscript. E.S. conceptualized and supervised the project.

