## Supplemental Information for "Epigenetic impairment and blunted transcriptional response to *Mycobacterium tuberculosis* of alveolar macrophages from persons living with HIV"

**Affiliations:**
[1]Department of Biochemistry, Faculty of Medicine, McGill University, Montréal, QC, Canada.

[2]Program in Infectious Diseases and Global Health, The Research Institute of the McGill University Health Centre, Montréal, QC, Canada.

[3]McGill International TB Centre, Department of Medicine, Faculty of Medicine, McGill University, Montréal, QC, Canada.

[4]Translational Research in Respiratory Diseases Program, The Research Institute of the McGill University Health Centre, QC, Montréal, Canada.

[5]Institute for Bio-computation and Physics of Complex Systems BIFI, Department of Theoretical Physics, University of Zaragoza, Zaragoza, Spain.

[6]Department of Medicine, Genetic Section, University of Chicago, Chicago, IL, USA.

[7]CHU Sainte-Justine Hospital Research Centre, Montréal, QC, Canada.

[8]Canadian Centre for Computational Genomics, McGill University and Genome Quebec Innovation Center, Montréal, QC, Canada.

[9]Chronic Viral Illnesses Service and Division of Hematology, McGill University Health Centre, Montréal, QC, Canada.

[10]Division of Clinical Infectious Diseases, Research Center Borstel, Borstel, Germany.

[11]International Health/Infectious Diseases, University of Lübeck, German Center for Infection Research (DZIF), Partner Site Hamburg-Lübeck-Borstel, Lübeck, Germany.

‡ Share first authorship
† Share senior authorship

**Supplementary Materials**

#### Materials and Methods

*RNA sequencing and raw data processing*

We generated 100 RNA-seq libraries encompassing pairs of baseline and *Mtb*-challenged samples for 16 control (HC), 14 PrEP and 20 PLWH subjects. Indexed libraries were combined in four to a maximum of 12 libraries per pool, with library pair per subject on the same pool, and sequenced as 100 bp single-end reads with an expected throughput of 25 million reads per library. Raw fastq files were quality-checked with FastQC (v0.11.8) [1], RSeQC (v2.6.1) [2], and subjected to trimming with cutadapt (v2.6) [3] to remove leftover adaptors and low-quality bases ($\leq$ 20 Phred score) allowing for a maximum read size of 99bp and a minimum of 75bp. Sequences were then aligned to the human reference genome GRCh38.p13 (ENSEMBL v99) [4] with STAR (v2.7.3a) [5]. Aligned BAM files were input to Salmon version 1.1.0 [6] for expression quantification.

*RNA expression matrix and gene filtering*

Text files containing estimated counts and transcript per million (TPM) values for 60,676 annotated features, for each subject, were used to create a gene level expression matrix in R (v3.6.1) [7] using tximport (v1.12.3) [8] and biomaRt (2.40.5) [9]. We adjusted the estimated counts by gene length and abundance (TPM) employing tximport's method "lengthScaledTPM" as discussed by Love *et al.* [10]. Next, we filtered out genes with estimated counts $\leq$ 10 in more than 70 libraries and ran edgeR [11] function "calcNormFactors" to generate scaling normalization factors with the TMM method [12]. Downstream analyses were focused on protein-coding genes with $\geq$ 100 estimated counts in at least 11 HC or 10 PrEP or 14 PLWH libraries, resulting in 10,362 genes for differential gene expression analyses. The gene matrix limited to the testable genes and the TMM scaling factors were input in limma [13] voom [14] (v3.40.6) to create log2-CPM normalized expression matrices and associated weights.

*ATACseq and ChIPseq data processing*

We profiled chromatin accessibility and H3k27 acetylation for non-stimulated and *Mtb* challenged AMs from 44 subjects and 16 subjects, respectively. Libraries were sequenced as 100bp paired end reads for ATACseq and 100bp single end reads for ChIPseq. Nextera adaptors and low quality reads were removed with TrimGalore v0.6.5 [3]. Reads were then aligned to both human (hg38) and *Mtb* (H37rv) genomes using BWA v0.7.17 default parameters [15]. PICARD v2.18.9 was used to mark duplicates and to assess the fragment length distribution. Reads aligning to the mitochondrial genome were removed with samtools v1.9 [16]. Next, alignmentSieve v3.3.2 was used to: (i) select unique paired-end reads (ATACseq) or unique single end reads (ChIPseq) using the --samFlagExclude 1804 and --samFlagExclude 1796 flags, respectively; (ii) remove ENCODE hg38 blacklisted regions [17]; and (iii) to extract fragments between 40 – 2000bp in length [18]. After quality control with MultiQC v1.8 [19], ATACseq libraries from two subjects yielded less than 20 million unique reads, and one library deviated from the periodic nucleosome pattern for ATACseq. Both the non-stimulated and *Mtb* challenged libraries of subjects failing QC were removed from further analysis.

*Peak annotation*

We integrated two approaches to annotate targeted genes for the 53,040 ATACseq tested peaks and 61,535 H3K27ac regions from ChIPseq. First, we assigned peaks based on distance < 5kb from a gene transcriptional start site (TSS) using ChIPSeeker v1.24 [20]. Next, we intersected our peak set with the GeneHancer 2017 dataset using mergeByOverlaps from IRange package v2.22.2 [21, 22]. GeneHancer includes a collection of regulatory regions assigned to genes based on the integration of multiple genome-wide regulatory databases [21]. By combining the TSS and GeneHancer approaches ~80% of the tested peaks were assigned to at least one gene (Figure S5).

*Chromatin accessibility and H3K27ac filtering, and library normalization*

We performed a count-based quantification of accessible chromatin and H3k27 acetylation as shown in [23]. Briefly, for each library MACS2 v2.2.6 callpeak was used with --call-summits in BAMPE mode (ATACseq) or with --shift -37 and --extsize 73 flags (ChIPseq) to identify regions of open chromatin or H3k27 acetylation. Next, the summit of each peak was extended 250bp in both directions resulting in fixed-width peaks of exactly 501bp. We merged fixed-width peaks overlapping in at least two samples with bedtools v2.26 [24]. By restraining the overlap of accessible chromatin between samples to fixed-width peaks we avoid merging multiple independent regions into a single large peak. We identified 116,547 regions of accessible chromatin and 120,833 of H3k27 acetylation present in at least two libraries of each approach, respectively. Next, for each library, featureCounts v1.6.3 was used to count the number of unique fragments (ATACseq) or unique reads (ChIP) overlapping the targeted regions [25]. A matrix containing the individual counts for each approach was exported to R. Next, low count peaks were removed with edgeR's v3.30.3 filterByExpr using 25 counts as cut off and the quantification matrices were normalized with the upperquartile method implemented on calcNormFactors [11]. Counts were then transformed to log2 count per million (CPM) using limma's v3.44.3 voom and the resulting matrix was used for downstream analysis [13, 14] .

*Linear models*

Differences in chromatin accessibility, H3k27 acetylation and transcriptomic between non-stimulated and *Mtb* challenged AMs were analysed in paired designs. To create the models we employed the function model.matrix from stats package (v4.0.2) using the factors: sample group (HC, PrEP and PLWH), *Mtb* status (baseline and *Mtb* challenge) and subject IDs (to enable paired design models) as denoted by the following equation:

$$Log2(CPM)_f \sim \beta_0 + \sum_{i=1}^{n-1} \beta_i \cdot \chi_i + \beta_{(HC.Mtb)} \cdot \chi_{(HC.Mtb)} + \beta_{(PLWH.Mtb)} \cdot \chi_{(PLWH.Mtb)} + \beta_{(PrEP.Mtb)} \cdot \chi_{(PrEP.Mtb)} + \varepsilon$$

Log2(CPM) represents the depth-normalized log-quantifications with $f$ indicating each gene or peak tested. $\beta_o$ is the intercept term, representing log-quantification for an arbitrary non-challenged library acting as reference. The term $\beta_i$ represents the individual-wise difference between the non-challenged sample from the i-th individual and the reference sample. $\chi_i$ is thus a dummy variable marking the identity of the (n-1) individuals in the dataset, excluding the reference. The following three terms in the equation capture the group-specific *Mtb* challenge effects nested within each group. The $\beta$ value for each of these terms corresponds to the logFC for the *Mtb* challenge per group after accounting for the inter-individual variance at the baseline present in our datasets. For ATAC-seq we included two additional standardized technical covariates in the linear model: the total number of unique reads per library and Bioanalyser average fragment length.

*Differential gene expression, chromatin accessibility and H3k27 acetylation in response to Mtb.*

Differences in RNA expression, chromatin accessibility and H3k27 acetylation between non-stimulated and *Mtb* challenged AMs were tested in parallel with limma. After defining the linear models, we used makeContrasts to guide the calculation of i) group-specific *Mtb* effects and ii) differential *Mtb* responses between groups (for RNA-seq only). For ii, a) $\beta_{(PLWH.Mtb)} - \beta_{(HC.Mtb)}$ was the difference in the response to *Mtb* of PLWH minus HC, b) $\beta_{(PrEP.Mtb)} - \beta_{(HC.Mtb)}$ the response difference of PrEP minus HC and c) $\beta_{(PLWH.Mtb)} - \beta_{(PrEP.Mtb)}$ the difference between PLWH minus PrEP. Of note, for ATACseq, we removed flowcell batch effects with ComBat as the paired nature of our approach prevented the inclusion of this covariate in a linear model [26, 27]. We then applied voomMod from cbcbSEQ to recalculate sample weights after batch correction. Next, we fitted the linear models with limma's function lmFit, derived coefficients for the contrasts defined via makeContrasts with ebayes [13] and extracted results with topTable. For ATAC and ChIP-seq we estimated false discovery rate (FDR) with the qvalue package v2.20.0 and considered peaks as significant if FDR $< 0.05$ and absolute log fold-change (logFC) $> 0.2$. For RNA-seq we employed the FDR procedure implemented in stageR v 1.6.0 [28] and considered genes as differentially expressed if absolute logFC $\geq 0.2$ and stageR *p*-val (stgr.p) $\leq$ 0.05.

*Gene enrichment analysis*

Pathways and gene ontology (GO) enrichment analysis were performed with clusterProfiler package version 3.16.0 and ReactomePA version 1.32.0, using differentially expressed genes (DEGs), genes assigned to DOCs or differentially acetylated (DAc) regions. Three different databases and functions were used for the enrichment analysis, KEGG was tested with enrichKEGG function, Reactome with enrichPathway and GO biological process was tested via enrichGO [29]. GO terms and pathways that had less than five assigned genes were excluded. Next, we merged the results from the three approaches and used the Benjamini-Hochberg's method to estimate the FDR. For DEGs enrichment we used FDR ≤ 0.1 while the cutoff for ATAC and ChIP-seq was FDR ≤ 0.05.

*Data visualization*

To visualize region of accessible chromatin as genomic tracks we applied a scaling factor based on edgeR's upperquartile normalization method. Of note, as the ATACseq and ChIPseq quantifications considered only fragments/reads in peaks (FrIP), normalizing genomic tracks by total library depth does not correspond to the quantification analysed by the linear models. To address that, a scaling factor per sample was calculated for genomic tracks as the reciprocal of $(\frac{Upperquartile\ norm\ x\ Total\ FrIP}{1000000})$. DeepTool's bamCoverage was used to produce scaled BigWig and bedGraphs from analysed BAM files [30]. We used SparK to calculate the mean coverage and standard deviation per base-pair per group for the genomic track plots [31]. Volcano and rankPlots were produced by adapting tools from the MAGeCKFlute package [32]. For the boxplots we used the residual log2CPM after regressing out covariates included in the linear models with removeBatchEffect from limma [13]. Density, Manhattan, bar and dot plots were produced using ggplot2 [33].

*Motif enrichment and footprint analysis*

The findMotifsGenome function (HOMER v4.11) was used to assess enrichment of TF motifs in DOCs and DAc regions versus all other tested peaks as background [34]. We used HINT-ATAC from RGT v0.13.0 to identify TF footprints and to estimate TF activity over footprints located in DOCs and DAc regions [35]. The analysis for differential TF activity over footprint compared the average depth of footprints encompassing motifs of a given TF between paired non-stimulated and *Mtb* challenged AM. This approach produced an activity score per TF with a corresponding $p$ value for each subject t [35]. TF containing an average of less than 50 footprints across samples were excluded from the analysis. We used a meta-analysis to combine $p$ values from each paired comparison using the sum of Z Stouffer's method from metap v1.4 package and applied a q-value FDR correction over meta-analysed $p$ values. TF with FDR < 0.05 and activity score > 0.005. We used a hypergeometric test to estimate the enrichment of IRF9 and ZNF684 active footprints in the TSS of DEGs and GO/Pathway analysis.

### Supplementary Figures

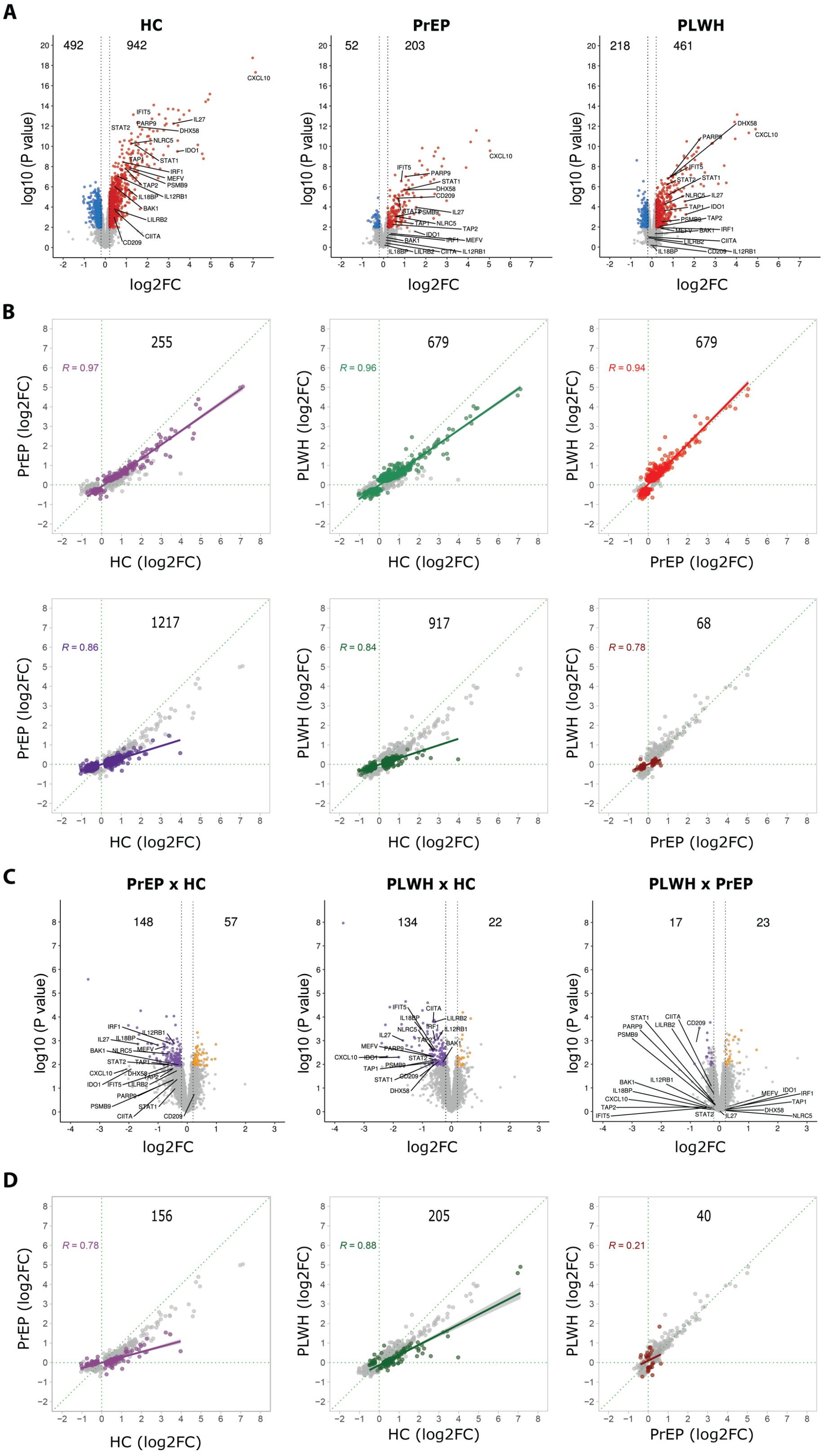

**Fig. S1. mRNA profile in response to *Mtb*.**

**(A)** Volcano plots for healthy controls (HC), subjects on pre-exposure prophylaxis (PrEP), and persons living with HIV receiving antiretroviral treatment (PLWH). Differentially expressed genes (DEG) in response to *Mtb* are displayed as function of log2FC (x-axis) and negative log10 unadjusted *P*-value (y-axis). Each dot corresponds to a single gene, vertical dashed lines indicate the logFC -0.2 and 0.2 thresholds. Red and blue dots represent genes significant at stageR FDR ≤ 5% that were, respectively, up- or down-regulated. Grey dots are genes that did not meet the log2FC thresholds (inside dashed lines) or that were not significantly differentially expressed (below coloured dots). Genes identified by symbols were significant DE for the HC response to *Mtb*, were significant for the PLWH vs HC differential gene response (panel **C**, middle), were in significant GO-terms/pathway for the PLWH vs HC contrast, and had significant DOCs. These genes are prototypical examples for the diminished response by alveolar macrophages (AM) from PLWH to *Mtb*. **(B)** Scatterplots for log2FC from *Mtb* challenge effect. log2FC for two groups are plotted against each other with the group on the x-axis considered the reference group. Grey dots represent the union of DEG from both groups. Top panel coloured dots indicate DEGs for the y-axis group (counts for DEG y-axis group are shown). Pearson correlations (R value indicated) were calculated based on the coloured dots irrespective if the corresponding gene in the x-axis group reached significance for DEG. Bottom panels contain the same data and order as the top ones, however coloured dots now represent DEGs for the x-axis group. As above, correlations were calculated for coloured log2FC pairings. These plots illustrate the overall blunted transcriptional response to *Mtb* by AM from PLWH and PrEP subjects. **(C)** Volcano plots for *Mtb* response differences between groups (PrEP vs HC; PLWH vs HC; PLWH vs PrEP). Purple dots represent significant genes with diminished log2FC (lower response) for PLWH or PrEP in contrast to HC subjects. Yellow dots represent significant genes with increased log2FC (higher response) for the non-reference groups. Grey dots depict non-significant genes for the response differences presented. Annotated genes are the same as described in panel **A**. **(D)** Genes differentially triggered between groups. As in panel **B**, grey dots represent the union of DEG from both groups. Coloured dots indicate DEG detected by the interaction terms for the groups indicated on the x and y-axes. The counts of DEG for the group contrasts is indicated on top of each panel. The x-axis depicts the *Mtb* effect (log2FC) for the indicated reference group while the y-axis shows the log2FC of the *Mtb* effect for the contrasted group. The correlations show that the majority of PLWH vs HC and PrEP vs HC DE genes were more strongly induced in HC subjects.

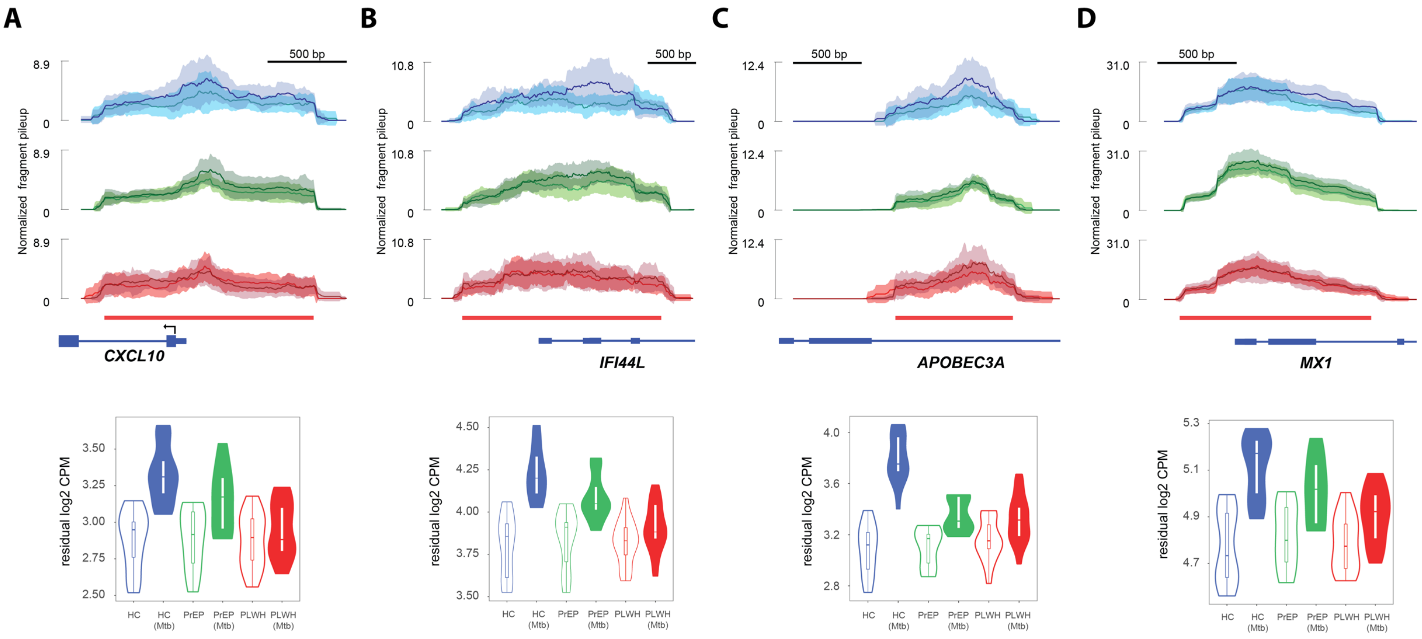


**Fig. S2. Changes in chromatin accessibility of DEG with high log2FC in response to *Mtb*.**

Chromatin accessibility for healthy controls (HC), subjects on pre-exposure prophylaxis (PrEP), and persons living with HIV receiving antiretroviral treatment (PLWH) is depicted for the transcriptional start site (TSS) of four DEG **(A)**, *CXCL10*, **(B)**, *IFI44L*, **(C)**, *APOBEC3A*, and **(D)**, *MX1*, that are among the genes displaying the highest log2FC transcriptional changes in response to *Mtb*. At the top, the mean normalized fragment pileup is plotted on the y-axis for HC, PrEP and PLWH groups and coloured in blue, green, and red, respectively. The lines indicate the mean accessibility for each condition in the group with a light shade representing non-infected AM and a darker shade the *Mtb*-challenged AM. The standard deviation of the means is shown by the corresponding shades for each condition. The red bar indicates the region shown in the quantification violin plots at the bottom. The residual log2 copy per million (CPM) for chromatin accessibility after removing inter-individual variability, is plotted for each group (HC, PrEP, and PLWH) and condition (*Mtb*-challenged or not) on the y-axis.

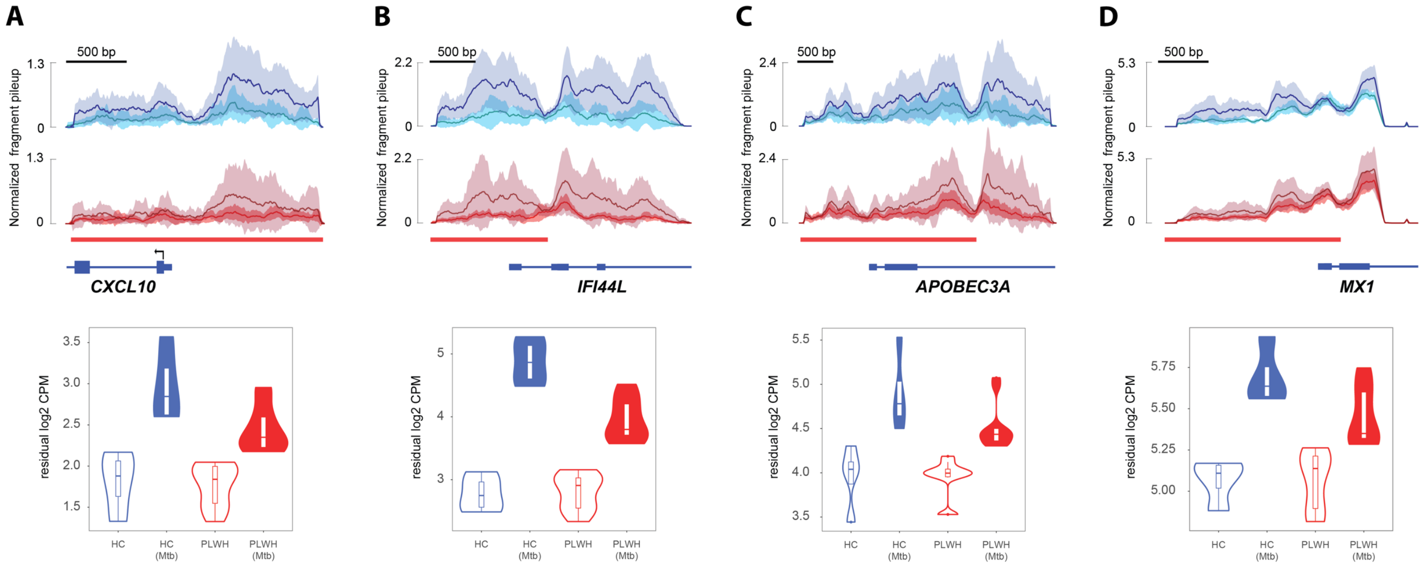


**Fig. S3. Changes in H3K27ac of DEG with high log2FC transcriptional changes in response to *Mtb*.**

H3K27ac for healthy controls (HC), and persons living with HIV receiving antiretroviral treatment (PLWH) is depicted for the TSS of four DEG **(A)**, *CXCL10*, **(B)**, *IFI44L*, **(C)**, *APOBEC3A*, and **(D)**, *MX1*, that are among the genes displaying the highest log2FC in response to *Mtb*. The mean normalized fragment pileup is plotted on the y-axis for HC and PLWH coloured in blue and red, respectively. The lines indicate the mean accessibility for each condition in the group with a light shading representing non-infected alveolar macrophages (AM) and a darker shading identifying *Mtb*-challenged AM. The standard deviation of the means is shown by the corresponding shades for each condition.

#### Supplementary Tables

#### Table S1. Differential gene expression and term enrichment test summary.

| Test | Contrast | DEGs | # Genes FDR ≤ 0.05 | % Genes in combined-terms[a] | # combined-terms FDR ≤ 0.05[b] | KEGG | | Reactome | | GO BP | |
|---|---|---|---|---|---|---|---|---|---|---|---|
| | | | | | | FDR 5% | Tested | FDR 5% | Tested | FDR 5% | Tested |
| *Mtb* challenge effect | HC + *Mtb* vs HC | Combined | 1434 | 76.56 | 1445 | 77 | 219 | 209 | 477 | 1159 | 2885 |
| | | Up-regulated | 942 | 81.1 | 1452 | 83 | 168 | 223 | 373 | 1146 | 2361 |
| | | Down-regulated | 492 | 67.88 | 90 | 7 | 48 | 7 | 68 | 76 | 989 |
| | PLWH + *Mtb* vs PLWH | Combined | 679 | 74.22 | 845 | 59 | 132 | 82 | 191 | 704 | 1776 |
| | | Up-regulated | 461 | 80.91 | 851 | 57 | 99 | 64 | 132 | 730 | 1446 |
| | | Down-regulated | 218 | 60.09 | 16 | 0 | 8 | 2 | 20 | 14 | 390 |
| | PrEP + *Mtb* vs PrEP | Combined | 255 | 68.23 | 254 | 25 | 32 | 17 | 48 | 212 | 623 |
| | | Up-regulated | 203 | 72.41 | 286 | 26 | 29 | 17 | 32 | 243 | 515 |
| | | Down-regulated | 52 | 51.92 | 17 | 1 | 1 | 1 | 1 | 15 | 19 |
| *Mtb* response differences | PLWH vs HC | Combined | 156 | 71.79 | 262 | 29 | 33 | 28 | 36 | 205 | 355 |
| | | Higher response[c] | 22 | 50 | 0 | 0 | 0 | 0 | 0 | 0 | 0 |
| | | Lower response[d] | 134 | 75.37 | 261 | 31 | 33 | 28 | 36 | 205 | 355 |
| | PrEP vs HC | Combined | 205 | 70.24 | 373 | 30 | 41 | 30 | 41 | 313 | 573 |
| | | Higher response | 57 | 56.14 | 6 | 0 | 0 | 0 | 0 | 6 | 6 |
| | | Lower response | 148 | 148 | 363 | 29 | 34 | 30 | 34 | 304 | 434 |
| | PLWH vs PrEP | Combined | 40 | 25 | 0 | 0 | 0 | 0 | 0 | 0 | 0 |
| | | Higher response | 23 | 26.08 | 5 | 0 | 0 | 0 | 0 | 5 | 5 |
| | | Lower response | 17 | 23.52 | 0 | 0 | 0 | 0 | 0 | 0 | 0 |

[a] To derive gene percentage in significant GO terms/pathways, we considered the results from "Combined DEGs" enrichment testing.

[b] Only terms with at least 5 assigned genes were considered.

[c,d] Refers to positive and negative log2FC results, respectively, from each interaction test.

#### Table S2. ATAC-seq and H3k27ac differential accessibility/mark and term enrichment test summary.

| Approach | Contrast | Peaks | # Regions FDR < 0.05 | Regions assigned to genes | # genes[a] | # genes in terms | KEGG | | Reactome | | GO BP | |
|---|---|---|---|---|---|---|---|---|---|---|---|---|
| | | | | | | | FDR 5% | Tested | FDR 5% | Tested | FDR 5% | Tested |
| **ATACseq** | HC + *Mtb* vs HC | Combined | 12369 | 8425 (68.1%) | 16085 | 9850 (61.2%) | 63 | 323 | 41 | 1395 | 620 | 6348 |
| | | Open | 8389 | 5355 (63.8%) | 11562 | 7279 (63.0%) | 42 | 316 | 42 | 1338 | 461 | 6001 |
| | | Close | 3971 | 3070 (77.3%) | 6430 | 3872 (60.2%) | 3 | 309 | 5 | 1047 | 23 | 4549 |
| **H3K27ac** | HC + *Mtb* vs HC | Combined | 198 | 148 (74.7%) | 514 | 353 (68.8%) | 19 | 68 | 21 | 109 | 127 | 729 |
| | | More acetylated | 173 | 131 (75.7%) | 481 | 332 (69.0%) | 20 | 67 | 23 | 106 | 127 | 683 |
| | | Less acetylated | 25 | 17 (68.0%) | 33 | - | - | - | - | - | - | - |

[a] Genes can be assigned to both open and closed differential open chromatin.

#### Table S3. Enrichment of IRF9 and ZNF684 footprints in the promoter of DEGs.

| | | DEG in HC + *Mtb* | | | |
|---|---|---|---|---|---|
| | | Yes | No | **Total** | |
| **IRF9 active footprint in the gene TSS** | Yes | 65 | 70 | 135 | $p = 7.9 \times 10^{-23}$ |
| | No | 1369 | 8858 | 10227 | |
| **ZNF684 active footprint in the gene TSS** | Yes | 258 | 1047 | 1305 | $p = 5.3 \times 10^{-11}$ |
| | No | 1176 | 7881 | 9057 | |
| | **Total** | 1434 | 8928 | 10362 | |
